# Serum miRNA-based signature indicates radiation exposure and dose in humans: a multicenter diagnostic biomarker study

**DOI:** 10.1101/2021.08.23.21262229

**Authors:** Zuzanna Nowicka, Bartłomiej Tomasik, David Kozono, Konrad Stawiski, Thomas Johnson, Daphne Haas-Kogan, Marek Ussowicz, Dipanjan Chowdhury, Wojciech Fendler

**Affiliations:** Department of Biostatistics and Translational Medicine, Medical University of Lodz, Poland; Department of Radiation Oncology, Dana-Farber Cancer Institute, Boston, MA, USA; Brigham and Women’s Hospital/Dana-Farber Cancer Institute, Harvard Medical School, Boston, MA, USA; Department of Pediatric Hematology, Oncology and Bone Marrow Transplantation, Wroclaw Medical University, Poland

## Abstract

Mouse and non-human primate models showed that serum miRNAs may be used to predict the biological impact of radiation doses. We hypothesized that these results can be translated to humans treated with total body irradiation (TBI), and that miRNAs may be used as clinically feasible biodosimeters. To test this hypothesis, serial serum samples were obtained from 25 patients who underwent allogeneic stem-cell transplantation and profiled for miRNA expression using next-generation sequencing. Circulating exosomes were extracted, their miRNA content sequenced and cross-referenced with the total miRNA fraction. Finally, miRNAs with diagnostic potential were quantified with qPCR and an artificial neural network model was created and validated on an independent group of 12 patients with samples drawn under the same protocol. Differential expression results were largely consistent with previous studies and allowed us to build an 8-miRNA-based model that showed AUC of 0.97 (95%CI 0.89-1.00) and validate it using qPCR in an independent validation set where it showed accuracy >91% for detecting exposure and 87.5% for differentiating between lethal and non-lethal doses. MiRNAs used in the model were miR-150-5p, miR-126-5p, miR-375, miR-215-5p, miR-144-5p, miR-122-5p, miR-320d and miR-10b-5p. Additionally, miRNAs with detectable expression in this and two prior animal sets almost perfectly separated the irradiated from non-irradiated samples in mice, macaques and humans, validating the miRNAs as radiation-responsive through evolutionarily conserved transcriptional regulation mechanisms. We conclude that serum miRNAs reflect radiation exposure and dose for humans undergoing TBI and may be used as functional biodosimeters for precise identification of people exposed to clinically significant radiation doses.

## INTRODUCTION

Radiation exposure as a result of natural disaster or reactor accident is an ongoing threat that may potentially expose people to life-threatening doses of ionizing radiation to the entire body, akin to total body irradiation (TBI). In addition, a potential terrorist attack using an improvised nuclear device, or so-called dirty bomb, remains a worrisome scenario [1,2]. Regardless of the cause, such events will lead to exposure of many subjects to ionizing radiation in doses that will be unknown to emergency and medical personnel. The clinical effect for the particular human is the result of both the absorbed dose and individual’s radiosensitivity. Acute radiation syndrome (ARS) will develop in 10-50% of patients exposed to a dose of approximately 1 Gy [3]. There are three main clinical components of ARS: hematopoietic sub-syndrome (developing after exposure to 1-7 Gy within few weeks to 2 months), gastrointestinal sub-syndrome (after 8-12 Gy) and cerebrovascular sub-syndrome (developing after exposure to more than 12 Gy); the last two lead to death of a majority of exposed individuals [4]. Another potential measure used to gauge toxicity is the dose expected to cause death of 50% of an exposed population (LD50). For humans, the LD50 at 30 days (LD50/30) due to ionizing radiation exposure is approximately 2.5–4.5 Gy [5]. While it is possible to safeguard patients, rescue workers or the military by radioprotective agents given prior to exposure, the necessity of administering these medications within 24 hours prior to the event makes them impractical for the general public [6]. The proper time window for the administration of agents that mitigate radiation injury is also narrow and their efficacy varies greatly depending on the extent of the radiation dose received by an individual [7]. Therefore, in a widespread, high dose exposure scenario, only the proper triage of patients based on their likely radiation exposure will assure the delivery of proper care and maximize odds of survival. Taken into account that, as of now, despite numerous efforts [8], no biodosimeter has been approved by the U.S. Food and Drug Administration (FDA), there is an acute need for efficient biomarkers of high-dose radiation exposure.

MicroRNAs (miRNAs) are small noncoding RNAs present in body fluids such as serum and plasma. Their expression levels in these fluids have recently emerged as promising biomarkers for different pathological conditions [9,10]. Interestingly, several studies have associated levels of specific circulating miRNAs with various pathological conditions, including exposure to ionizing radiation [11,12]. The stability of serum miRNAs under various conditions and reproducible levels in individuals of the same species make these molecules compelling candidates for their use as noninvasive biomarkers [13]. A recent systematic review and meta-analysis showed that miRNAs may serve as robust biomarkers of radiation [14]. In addition, miRNA were reported to be cell- and tissue-specific and it has been postulated that circulating miRNA profiles may reflect the degree of tissue injury [15–17]. Several works, carried out predominantly on mice or non-human primates (NHPs), reported the discovery and validation of circulating miRNAs as biomarkers of radiation response [18–22]. Those studies showed also that serum/plasma miRNAs are evolutionarily conserved, which suggests that they could be also used as biomarkers in humans. Multiple studies have also shown that radiation affects the content of extracellular vesicles (exosomes) released both from cancer cells [23–25] as well as bystander stromal cells located within the irradiated field [26]. Furthermore, surface markers of the exosomes released by cells were reported to delineate both their origin as well as tissue targets [27]. The information transfer mediated by exosomal contents may serve as cell-to-cell communication used for the regulation of such fundamental phenomena like apoptosis evasion [28,29].

To address the question whether serum circulating miRNAs may predict the biological impact of lethal and non-lethal radiation doses in humans, we analyzed samples obtained from 25 prospectively recruited patients who underwent TBI as a component of conditioning prior to allogeneic stem-cell transplantation at two tertiary cancer centers. This scenario is the only existing proxy of a lethal dose exposure of humans if not followed by transplantation and, despite the potential interference of the underlying malignancy, is one that can be used to evaluate the impact of low or myeloablative doses of ionizing radiation. To provide a comprehensive description of the proposed biodosimeters, we have also assessed changes in radiation-induced exosomal cargo and performed bioinformatic analyses to identify the putative source of the radiation-induced miRNAs. Finally, we show signatures able to detect radiation exposure and differentiate between low and high dose in humans.

## RESULTS

### Radiation-induced miRNA expression changes in human serum

To identify miRNAs whose expression in human serum is altered by radiation exposure, i.e. potential biodosimeters, we obtained sera from 25 individuals undergoing TBI as a component of conditioning for allogenic hematopoietic stem cell transplantation (HSCT). Patients were recruited from two centers; the first group were children with mean age 10.6 ± 3.8 years and the second group were adults, with mean age 48.9 ± 16.5 years. TBI was delivered to a total dose of 12 Gy, 2 Gy/fx (Fig. 1A). Blood was drawn from all patients before TBI, after low dose exposure (2-4 Gy), and additionally from 9 patients after 12 Gy (high dose exposure). The high dose timepoint was included in the study design to assess dose-dependent patterns in the expression of radiation-induced miRNAs. miRNA expression was measured in sera by miRNA sequencing, yielding on average 723.7 ± 205.5 miRNAs with non-zero reads in each sample. Clinical characteristics of the participants are summarized in Table S1. After quality control, 22 sample pairs were retained for analysis in the low dose setting and 7 pairs in the high dose setting (Fig. S1, S2). Paired comparisons of pre- and post-irradiation samples revealed differentially expressed miRNAs, including 7 deregulated in both settings (Fig. 1B); all had consistent direction of change in the low and high-dose exposure settings.

**Figure 1.**
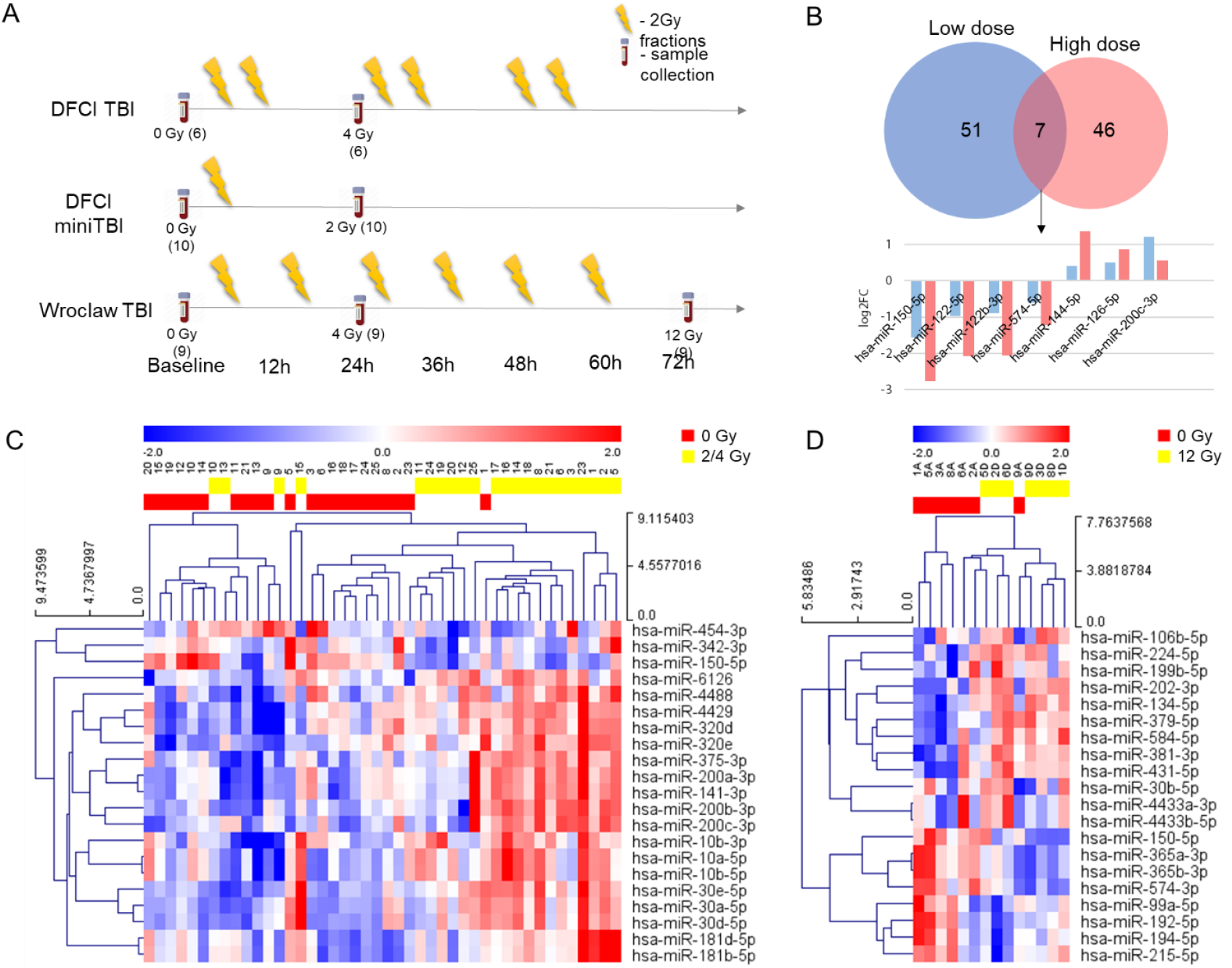
Total body radiation exposure impacts serum miRNA expression profile. (A) Sample collection. Numbers in parentheses denote the number of samples collected in given timepoint. (B) Venn diagram presenting overlap between miRNAs with p<0.05 in differential expression testing in the low- and high-dose comparisons. (C) Heatmap with hierarchical clustering of miRNAs differentially expressed in paired pre-irradiation samples and after 2/4 Gy in sera of patients undergoing TBI. Presented miRNAs had FDR<0.05 in differential expression testing of miRNAs with non-zero expression in at least one sample in a pair, in all pairs of samples. Red color represents high expression and blue represents low expression. (D) Heatmap with hierarchical clustering of miRNAs differentially expressed in paired pre-irradiation samples and after 12 Gy in sera of patients undergoing TBI. Presented miRNAs had p<0.015 in differential expression testing on miRNAs with non-zero expression in at least one sample in a pair, in all pairs of samples. Red color represents high expression and blue represents low expression.

Hierarchical clustering revealed clear separation of the samples from irradiated and non-irradiated patients based on the expression of top differentially expressed miRNAs – at FDR<0.05, 18 miRNAs were upregulated following low dose exposure and 3 were downregulated (Fig. 1C). No miRNAs were differentially expressed at FDR<0.05 in the high dose setting due to the exclusion of two out of nine high dose sample pairs after quality control and the resulting loss of statistical power. However, hierarchical clustering based on the expression of top 20 differentially expressed miRNAs showed very good separation of the irradiated and non-irradiated samples (Fig. 1D).

### High confidence miRNAs conserved between species and their putative tissue source

Having established the radiation-induced miRNA signatures in human serum, we aimed to select a set of miRNAs that could be used to design a clinically feasible, i.e. qPCR-based, test in humans. To this end, we cross-referenced the results from the present study with those from our earlier works in mouse and NHP models of radiation exposure (Fig. 2A). Because the doses of radiation exposure were not easily comparable between studies (2/4/12 Gy in humans, 2/6.5/8 Gy in mice and 5.8/6.5/7.2 Gy in macaques) we decided to merge the lists of differentially expressed miRNAs at p<0.05 for the low- and high-dose comparisons for humans and for mice. The result of this cross-species overlap is a list of eight miRNAs (Fig. 2B). Importantly, most overlapping miRNAs had consistent directions of change: three depleted (miR-215-5p, miR-150-5p, miR-122-5p) and three over-expressed (miR-30a-5p, miR-375 and miR-126-6p). For many miRNAs, the fold change was greater following high versus low dose radiation exposure (Fig. 2C).

**Figure 2.**
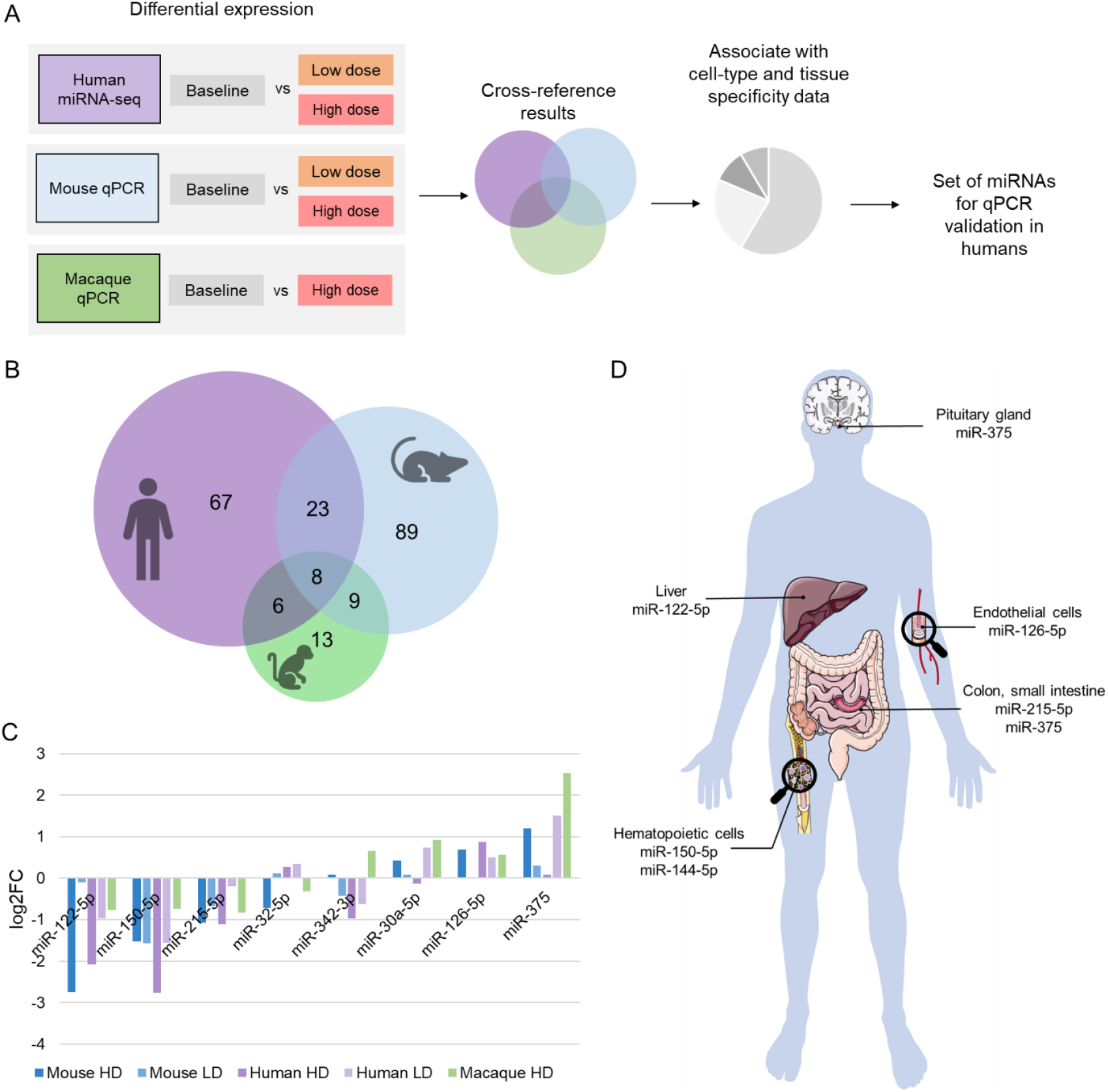
Selection of miRNAs for the qPCR-based study. (A) Schematic representation of the approach to identify candidate miRNAs for the model based on qPCR expression measurement. (B) Overlap between miRNAs with p<0.05 in the human (low and high dose lists combined), mouse (low and high dose lists combined) [reanalyzed from Acharya et al. 2015[18]], and macaque [reanalyzed from Fendler et al. 2017[19]] comparisons. (C) Radiation-induced miRNAs overlapping between the human, mouse and macaque datasets. Data are presented as log2 fold changes between the irradiated and non-irradiatied samples. LD – low dose, HD – high dose. (D) Putative tissue sources of miRNAs enriched (miR-181 family, miR-126, miR-375, miR-144-5p) or depleted (miR-150, miR-122, miR-215) in sera of patients undergoing TBI [based on the results from Landgraf et al. [17], Ludwig et al. [15] and de Rie et al. [16]

Besides the miRNAs with consistent direction of change in all three species, we noted other important overlaps among miRNA families (Table 1). Notably, besides miR-30a-5p, other miRNAs from the miR-30 family were frequently upregulated in sera: miR-30d-5p in human (both low and high-dose) and macaque; miR-30b-5p, miR-30c-5p and miR-30e-p in human and mouse. Also, miR-320c, miR-320d and miR320e were significantly upregulated in human serum, while miR-320-3p was upregulated in dose-dependent fashion in mouse (for the mouse study we used qPCR instead of miRNA sequencing and did not distinguish between miR-320 isoforms). miR-181a-5p, together with miR-181b-5p and miR-181d-5p, were significantly upregulated in human serum, while miR-181a-3p was upregulated in mouse serum. miR-200c-3p was significantly upregulated in human and mouse serum, while miR-200b-3p and miR-200a-3p were also upregulated in human and miR-200c-5p in mouse serum.

**Table 1.**
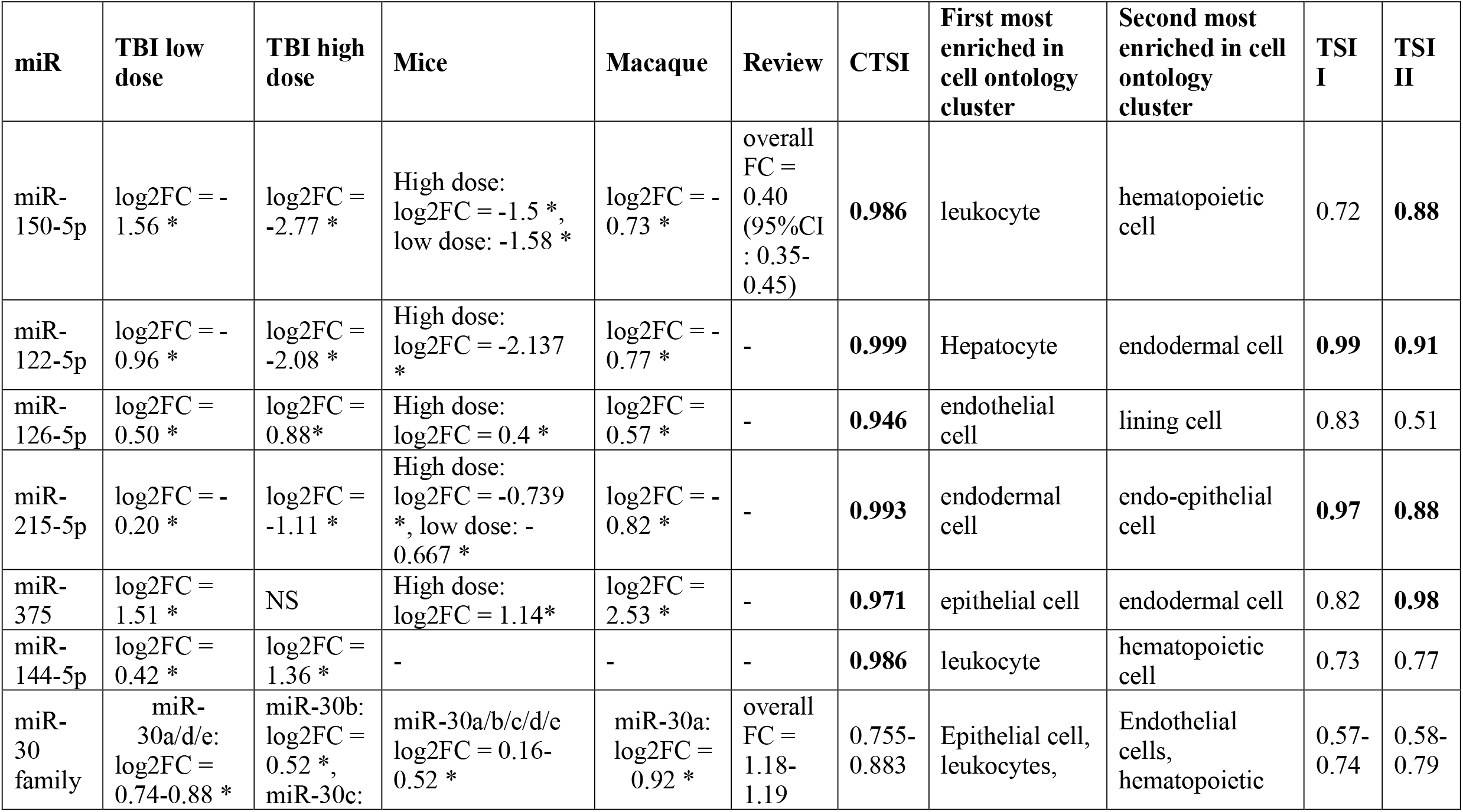
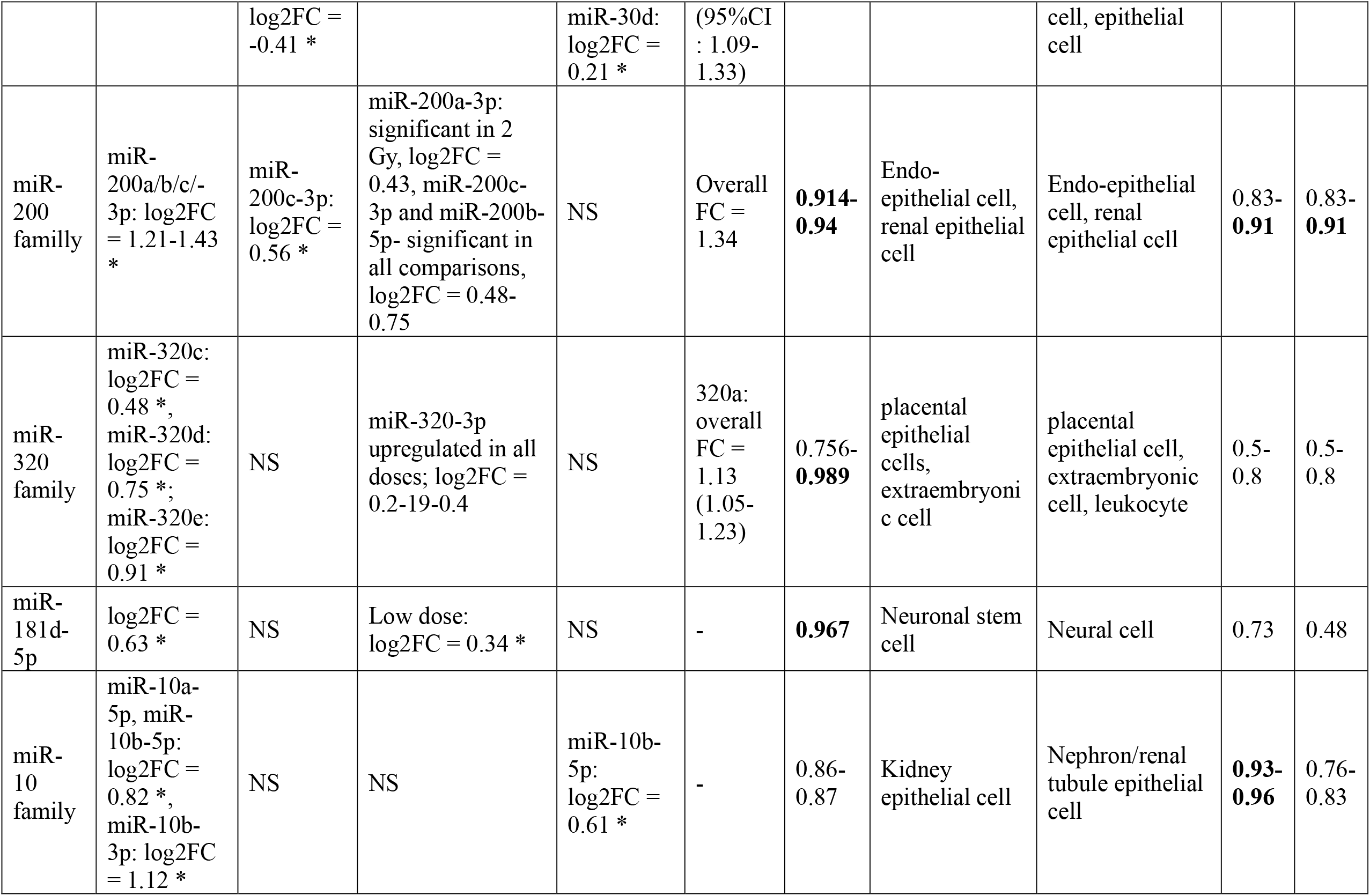
Summary information regarding miRNAs selected for the qPCR experiment. Values indicating cell-type specific (CTSI>0.9) and tissue-specific (TSI>0.85) expression patterns are presented in bold; significant expression changes are marked with an asterisk. NS – not significant. [miRNA expression data from mice – Acharya et al.[18]; 2015 miRNA expression data from macaques– Fendler et al. 2017[19]; Cell type specificity data – de Rie et al. [16]; Tissue specificity data – Ludwig et al. [15], Review – Malachowska et al. [14]]

| miR | TBI low dose | TBI high dose | Mice | Macaque | Review | CTSI | First most enriched in cell ontology cluster | Second most enriched in cell ontology cluster | TSI I | TSI II |
| --- | --- | --- | --- | --- | --- | --- | --- | --- | --- | --- |
| miR-150-5p | log2FC = -1.56 * | log2FC = -2.77 * | High dose: log2FC = -1.5 *, low dose: -1.58 * | log2FC = -0.73 * | overall FC = 0.40 (95%CI : 0.35-0.45) | **0.986** | leukocyte | hematopoietic cell | 0.72 | **0.88** |
| miR-122-5p | log2FC = -0.96 * | log2FC = -2.08 * | High dose: log2FC = -2.137 * | log2FC = -0.77 * | - | **0.999** | Hepatocyte | endodermal cell | **0.99** | **0.91** |
| miR-126-5p | log2FC = 0.50 * | log2FC = 0.88* | High dose: log2FC = 0.4 * | log2FC = 0.57 * | - | **0.946** | endothelial cell | lining cell | 0.83 | 0.51 |
| miR-215-5p | log2FC = -0.20 * | log2FC = -1.11 * | High dose: log2FC = -0.739 *, low dose: -0.667 * | log2FC = -0.82 * | - | **0.993** | endodermal cell | endo-epithelial cell | **0.97** | **0.88** |
| miR-375 | log2FC = 1.51 * | NS | High dose: log2FC = 1.14* | log2FC = 2.53 * | - | **0.971** | epithelial cell | endodermal cell | 0.82 | **0.98** |
| miR-144-5p | log2FC = 0.42 * | log2FC = 1.36 * | - | - | - | **0.986** | leukocyte | hematopoietic cell | 0.73 | 0.77 |
| miR-30 family | miR-30a/d/e: log2FC = 0.74-0.88 * | miR-30b: log2FC = 0.52 *, miR-30c: | miR-30a/b/c/d/e log2FC = 0.16-0.52 * | miR-30a: log2FC = 0.92 * | overall FC = 1.18-1.19 | 0.755-0.883 | Epithelial cell, leukocytes, | Endothelial cells, hematopoietic | 0.57-0.74 | 0.58-0.79 |
|  |  | log2FC =<br>-0.41 * |  | miR-30d:<br>log2FC =<br>0.21 * | (95%CI<br>: 1.09-1.33) |  |  | cell, epithelial<br>cell |  |  |
| miR-200<br>family | miR-200a/b/c/-<br>3p: log2FC<br>= 1.21-1.43<br>* | miR-200c-3p:<br>log2FC =<br>0.56 * | miR-200a-3p:<br>significant in 2<br>Gy, log2FC =<br>0.43, miR-200c-3p and miR-200b-5p- significant in<br>all comparisons,<br>log2FC = 0.48-0.75 | NS | Overall<br>FC =<br>1.34 | **0.914-**<br>**0.94** | Endo-epithelial cell,<br>renal epithelial<br>cell | Endo-epithelial<br>cell, renal<br>epithelial cell | 0.83-<br>**0.91** | 0.83-<br>**0.91** |
| miR-320<br>family | miR-320c:<br>log2FC =<br>0.48 *,<br>miR-320d:<br>log2FC =<br>0.75 *;<br>miR-320e:<br>log2FC =<br>0.91 * | NS | miR-320-3p<br>upregulated in all<br>doses; log2FC =<br>0.2-19-0.4 | NS | 320a:<br>overall<br>FC =<br>1.13<br>(1.05-1.23) | 0.756-<br>**0.989** | placental<br>epithelial<br>cells,<br>extraembryoni<br>c cell | placental<br>epithelial cell,<br>extraembryonic<br>cell, leukocyte | 0.5-0.8 | 0.5-0.8 |
| miR-181d-5p | log2FC =<br>0.63 * | NS | Low dose:<br>log2FC = 0.34 * | NS | - | **0.967** | Neuronal stem<br>cell | Neural cell | 0.73 | 0.48 |
| miR-10<br>family | miR-10a-5p, miR-10b-5p:<br>log2FC =<br>0.82 *,<br>miR-10b-3p: log2FC<br>= 1.12 * | NS | NS | miR-10b-5p:<br>log2FC =<br>0.61 * | - | 0.86-0.87 | Kidney<br>epithelial cell | Nephron/renal<br>tubule epithelial<br>cell | **0.93-**<br>**0.96** | 0.76-0.83 |

miRNA expression profiles are tissue-specific. We used data regarding tissue-specific miRNA expression in human [15] and mouse [17] and cell type-specific miRNA expression in human and mouse [16] to identify the putative source of the radiation-induced miRNAs in serum (Fig. 2D). We also accessed data from the study by de Rie et al. [16] to determine cell-type-specific miRNA expression in human and mouse. A summary of the findings from these three studies regarding the source of radiation-induced miRNAs selected for qPCR validation, together with information from this and previous animal studies on impact of radiation on their expression, is presented in Table 1.

### Radiation-induced miRNAs in serum-derived exosomes

Given the reports suggesting selective miRNA packaging into exosomes, we hypothesized that radiation-induced serum and exosome signatures might differ. To verify this speculation, we isolated exosomes from sera for 6 pairs of samples obtained in the high dose setting and performed miRNA sequencing of their contents (Fig. S4). At FDR<0.25, 7 miRNAs were upregulated and 4 miRNAs were downregulated (Fig. 3A). Overlap of miRNAs differentially expressed at p<0.05 in the low dose serum samples, high dose serum samples and exosomes revealed 4 shared miRNAs (miR-150-5p, miR-122-5p, miR-126-5p, miR-122b-3p; Fig. 3B) with consistent direction of change. However, the correlation between log2FCs of miRNAs in irradiated vs non-irradiated samples in exosomes and serum was weak (Spearman R=0.17, Fig. S5). miR-122-5p, miR-122-3p and miR-126-5p showed greater log2FC in exosomes compared to the serum high dose setting.

**Figure 3.**
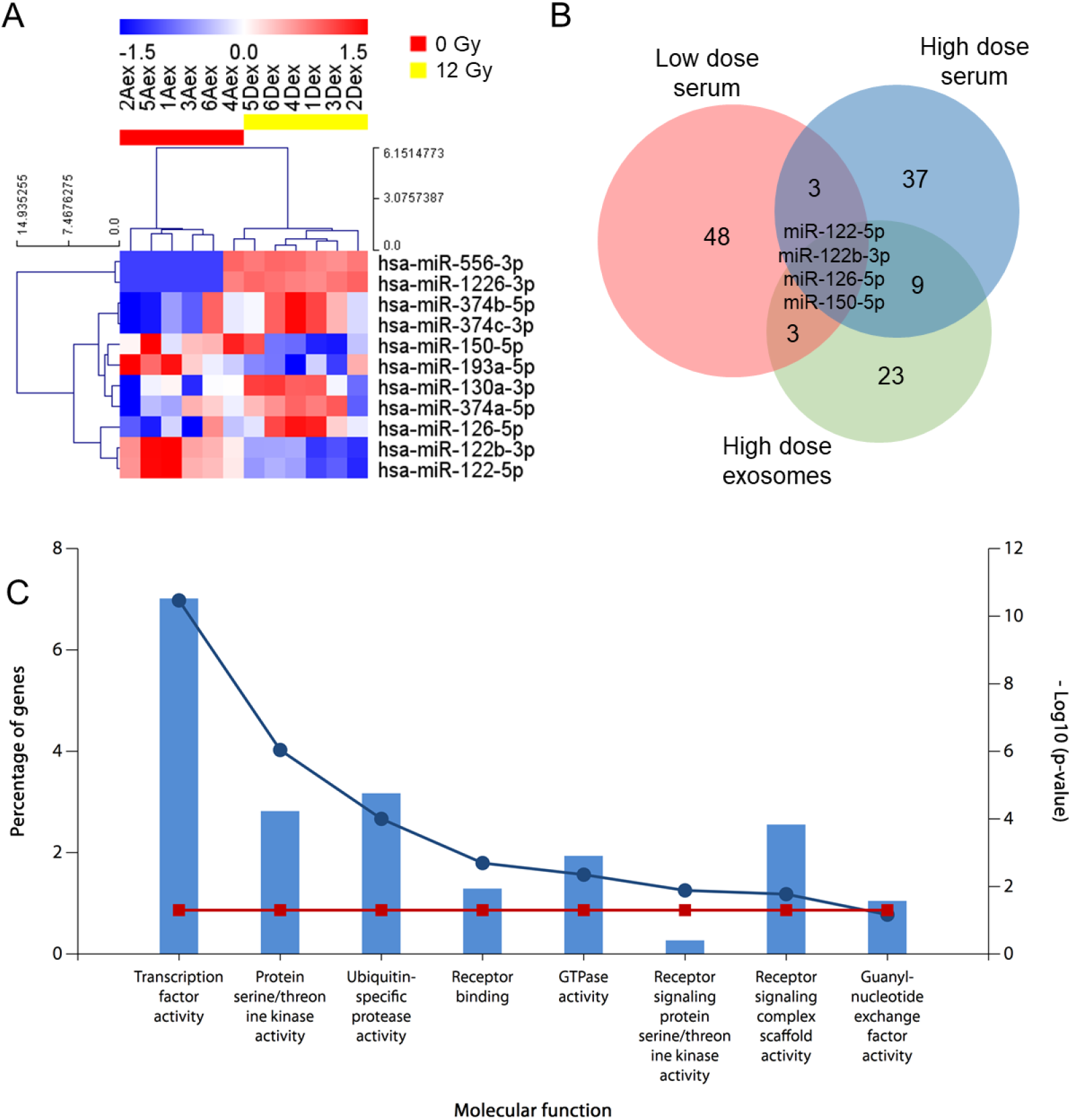
Changes in miRNA exosomal content after exposure to 12 Gy partly overlap with those observed in the serum low- and high-dose comparisons. (A) Heatmap with hierarchical clustering of miRNAs differentially expressed in paired pre-irradiation samples and after 12 Gy in exosomes of patients undergoing TBI. Presented miRNAs had FDR<0.25 in differential expression testing on miRNAs with non-zero expression in at least one sample in a pair, in all pairs of samples. Red color represents high expression and blue represents low expression. (B) Venn diagram presenting overlap between miRNAs with p<0.05 in differential expression testing in the serum low- and high-dose comparisons and in exosomes high-dose comparison. (C) Molecular functions associated with miRNAs differentially expressed (p<0.05) in exosomes isolated pre- and post-irradiation. Bar height shows percentage of genes and dots represent – log10(p value).

Since miRNA-containing exosomes were proposed to mediate cell-cell communication, especially in response to pathogenic stimuli[30], we hypothesized that exosomal miRNAs might be involved in the organism response to high dose irradiation. To address this question, i.e. to identify putative biological processes and pathways associated with miRNAs differentially expressed in exosomes before and after irradiation with 12 Gy, we used FunRich to perform functional enrichment analysis for all miRNAs significant at p<0.05 (Fig. 3C). Top significant molecular functions associated with those miRNAs include processes connected to transcription factor activity, receptor binding and receptor signaling.

### Evolutionarily conservation of radiation-responsive miRNAs

Having established the set of high-confidence radiation-responsive miRNAs in humans subjected to TBI, we assessed whether they can be used to identify a signature distinguishing the irradiated from non-irradiated samples using miRNA-sequencing data. We used logistic regression to create such a model in the low-dose setting (N=22 sample pairs). The resulting classifier was based on the expression of miR-150-5p, miR-126-5p and miR-375 and yielded an area under the receiver operating characteristic curve (AUC ROC) of 0.87 (95% CI: 0.76-0.98) in 4-fold cross validation (Fig. S6A). The Hosmer-Lemeshow test showed a good fit of the model (p=0.168).

For the high-dose detection scenario, we used the data of all available non-irradiated samples (N=22) and samples drawn after exposure to 12 Gy (N=9). A logistic regression model built using two of the three miRNAs from the low-dose signature (miR-150-5p and miR-126-6p; miR-375 was not significant in the high-dose comparison) yielded an AUC ROC of 0.85 (95% CI: 0.68-1.00) in 4-fold cross-validation (Fig. S6B). The model was well-fitted, with Hosmer Lemeshow test p=0.770. Therefore, the expression of miR-126-5p, miR-150-5p and miR-375 enabled efficient classification of the irradiated- and non-irradiated samples in both settings (Fig. S6C).

Then, to determine whether the observed differences in humans could be replicated in animal models without an underlying hematologic disease, we re-analyzed our previously published mice and macaque data[18,19] to build a logistic regression model using the same three miRNAs (miR-150-5p, miR-126-5p, miR-375). In the macaque samples, the resultant model maintained the efficacy that we observed in the human low-dose setting, classifying samples with AUC ROC=0.88 (95%CI: 0.78-0.98, Fig. S6D-E). In mice, the logistic regression model with the same input miRNAs allowed for perfect classification of all samples, both in the low- and high-dose setting (AUC ROC=1.00, 95%CI: 1.00-1.00 for both). The expression of miR-126-5p, miR-375 and miR-150-5p could therefore clearly distinguish irradiated and non-irradiated mouse samples (Fig. S6F). This confirmed that the identified miRNAs have the potential to robustly differentiate between different dose-exposure scenarios and their pattern of radiation-induced changes of serum expression are conserved across mammals. Having established that, we set upon defining an easy to use qPCR test for radiation exposure detection.

### Choice of endogenous normalizer miRNAs for qPCR validation

One vital aspect of translating the results from miRNA-seq based expression measurement in biofluids to applicable, quantitative RT-PCR (qPCR) tests was the identification of suitable internal reference miRNAs, i.e. normalizers. For this purpose, we used normiRazor – a tool that calculates stability scores for all potential 2- and 3-miRNA combinations in a dataset using several established algorithms and ranks miRNA combinations according to their stability. Since it has been proven that average expression of 2 miRNAs is in most cases statistically better in terms of stability, this approach is superior to the use of single reference miRNA. The highest-ranking pair identified for the miRNA-seq experiment was miR-27a-3p and let-7a-5p. Both miRNAs had high expression in all samples and were not significantly altered after irradiation, either with low or high dose (Fig. S7C). Their expression was also not altered in circulating exosomes after irradiation with high dose.

### qPCR validation of radiation-inducible miRNAs in human sera

To verify if the expression of selected miRNAs (listed in Table 1) normalized to endogenous reference can be used to predict radiation exposure and differentiate between low and high radiation dose, we used samples from the same patient cohort treated with TBI: 25 samples drawn at baseline, 15 samples after 4 Gy and 9 samples after 12 Gy (Fig. 4A). Additionally, we collected 9 samples 12 hours after the first 2 Gy fraction to determine if changes in serum miRNA expression can be detected in such short time period after radiation exposure. miRNA levels were normalized to the mean of miR-27a-3p and let-7a-5p.

**Figure 4.**
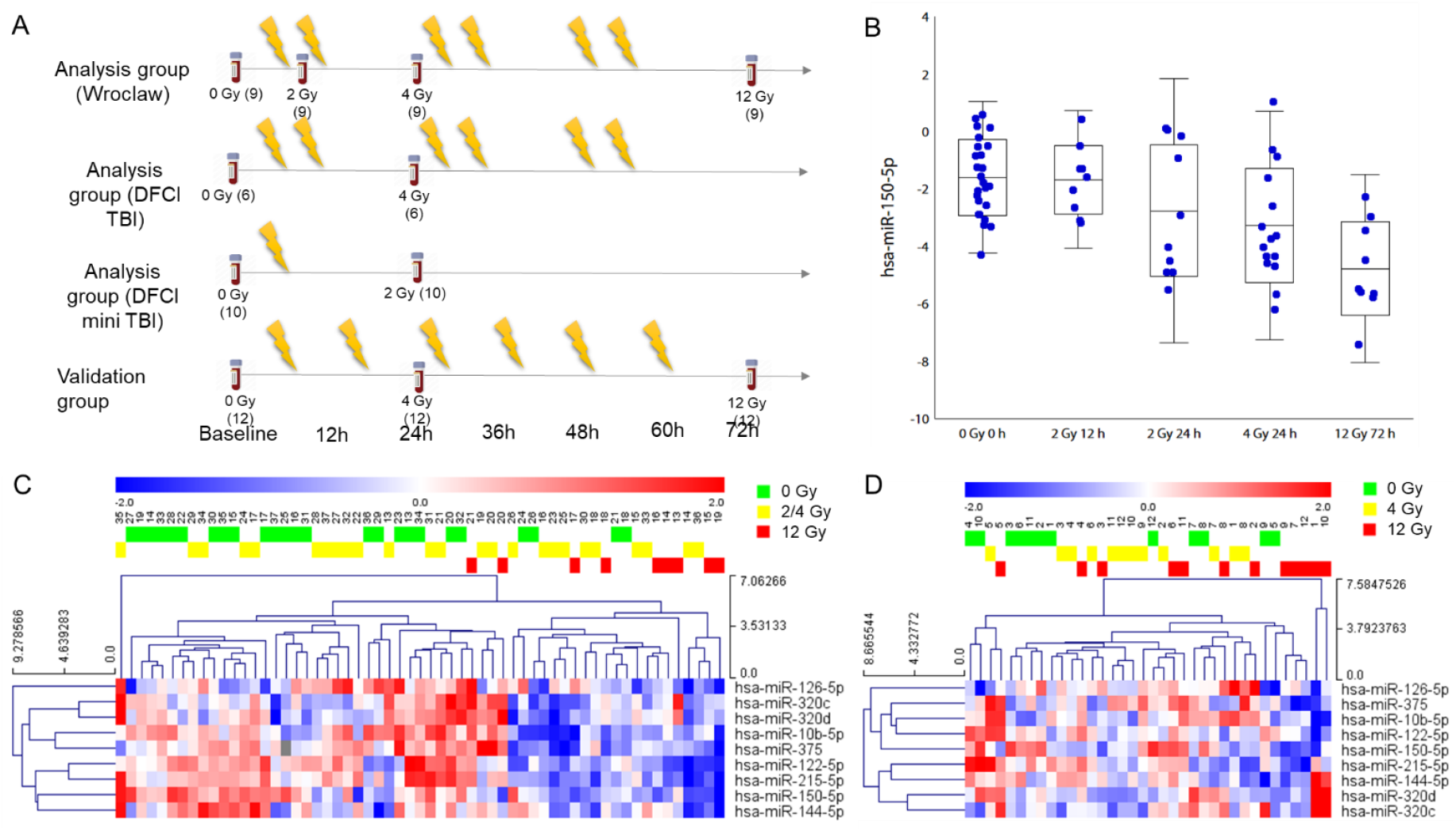
Development of a radiation biodosimeter using miRNA quantification in serum by qPCR. (A) Sample collection protocol. Numbers in parentheses denote the number of samples collected in given timepoint. (B) Expression of miR-150-5p normalized to mean expression of miR-27a-3p and let-7a-5p in samples collected at different timepoints before irradiation and after irradiation with 2, 4 or 12 Gy in the analysis cohort. (C) Heatmap with hierarchical clustering of selected miRNAs in sera of patients undergoing TBI from the analysis cohort. Red color represents high expression and blue represents low expression. (D) Heatmap with hierarchical clustering of miRNAs from Fig. 4C in sera of patients undergoing TBI from the validation cohort. Red color represents high expression and blue represents low expression.

None of the miRNAs expression changed significantly 12 hours after exposure to 2 Gy; the same was also true for miR-150-5p, which had lowest p value in baseline – 4 Gy and baseline – 12 Gy comparisons (Fig. 4B). Based on this result we excluded samples drawn 12 hours after exposure to 2 Gy from the low dose set. Notably, miR-150-5p expression kinetics in human samples was different from that observed in mice (Fig. S8).

Expression patterns distinguished samples collected at baseline and after low- and high-dose radiation exposure. Only one miRNA (miR-150-5p) was significantly downregulated at p<0.05 in the low dose setting, while miR-150-5p, miR-122-5p, miR-215-5p and miRNAs from the miR-30 family were downregulated in the high dose setting. In previous experiments, miRNAs from the miR-30 family were mostly upregulated after radiation exposure. Since miR-30 family shows extremely high sequence similarity, the discrepancy might arise from mismatches in the sequences mapping. We decided to exclude those miRNAs from further analysis, along with miRNAs with 10 or more missing values, leaving 9 miRNAs (Fig. 4C). Expression of the same miRNAs in the sera of 12 patients from the validation patients is presented in Fig. 4D. Clinical data of the patients from validation cohort are presented in Table S2.

Interestingly, a set of 3 miRNAs quantified by qPCR in all of our previous experiments clearly visually distinguished irradiated from non-irradiated samples in the human analysis (Fig. 5A) and validation cohort (Fig. 5B), in two macaque cohorts from the study by Fendler et al.[19] (Fig. 5C,D) and in mouse cohort from the study by Acharya et al.[18] (Fig. 5E).

**Figure 5.**
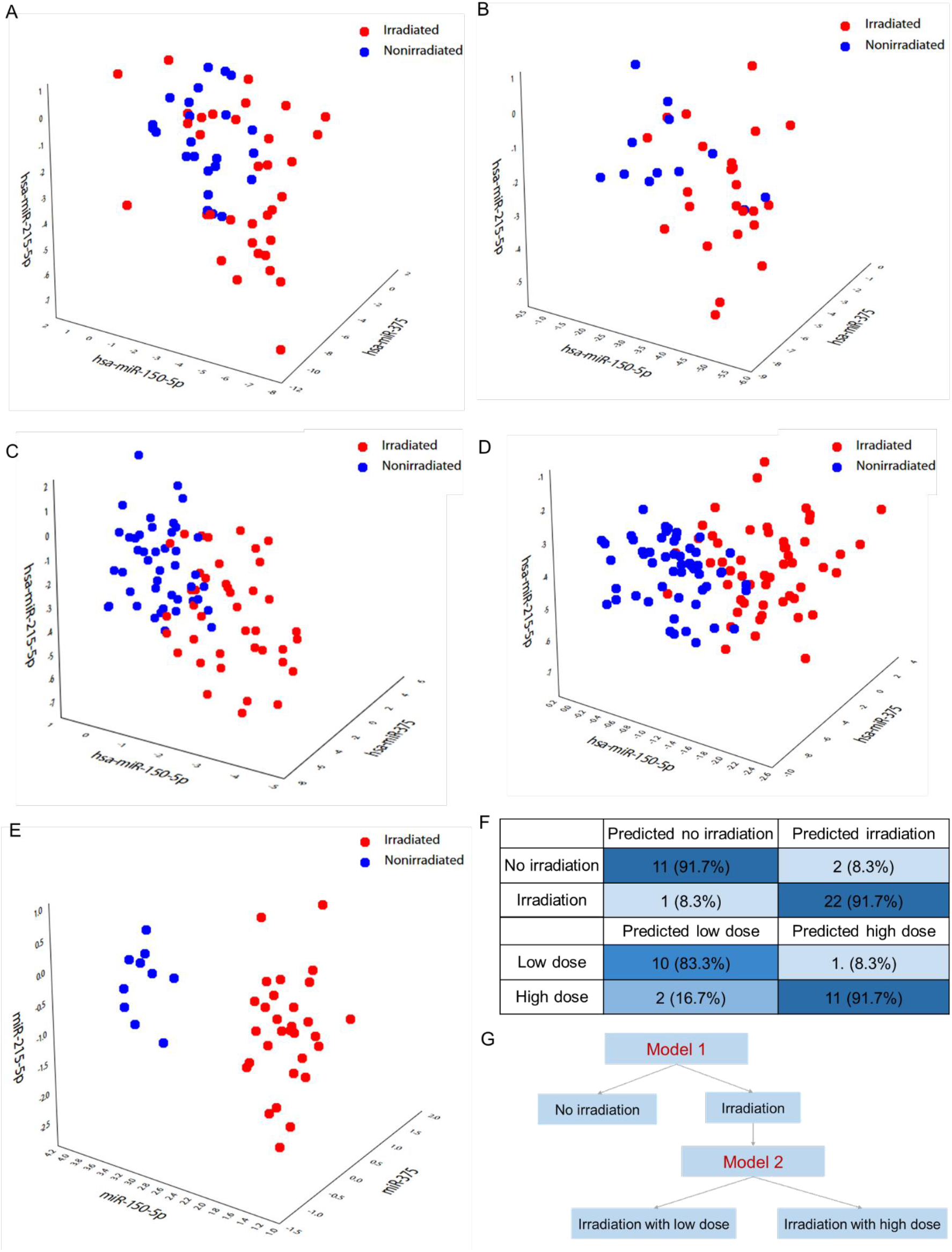
miRNA expression clearly separates irradiated from non-irradiated samples and allows to build neural network-based classification models. (A) Scatterplot of the expression of miR-215-5p, miR-150-5p and miR-375 in the analysis cohort. (B) Scatterplot of the expression of miR-215-5p, miR-150-5p and miR-375 in the validation cohort. (C) Scatterplot of the expression of miR-215-5p, miR-150-5p and miR-375 in the first macaque cohort from Fendler et al. (2017) and (D) in the macaque validation cohort from Fendler et al. (2017) (E) Scatterplot of the expression of miR-215-5p, miR-150-5p and miR-375 in the mice cohort from Acharya et al. (2015). (F) Prediction accuracy of neural network-based models to predict irradiation vs no irradiation and high vs low dose in the validation cohort. (G) Nine-miRNA diagnostic protocol for the prediction of irradiation and radiation dose. Model 1 is based on the expression of miR-10b-5p, miR-122-5p, miR-126-5p, miR-144-5p, miR-150-5p, miR-215-5p, miR-320d and miR-375 normalized to miR-27a-3p and let-7a-5p. Model 2 is based on the expression of miR-122-5p, miR-126-5p, miR-144-5p, miR-150-5p, miR-215-5p, miR-320d and miR-375 normalized to miR-27a-3p and let-7a-5p.

### Neural networks-based approach to detect radiation exposure and differentiate between low and high dose irradiation in humans

To achieve the best classification performance, we built neural network-based models as done in our previous work on circulating miRNA biomarkers[31]. Two networks were designed to work in a two-tier approach, to 1) identify samples from patients irradiated with any dose vs baseline samples based on serum miRNA expression measured by qPCR and 2) to differentiate between irradiation with low and high dose. To achieve sufficient sample size to divide the samples into the training and test set, we used all baseline, low and high dose samples, both paired and unpaired, to build the models; samples from the 12 patients from the validation cohort served as the validation set. Furthermore, we used Synthetic Minority Oversampling Technique (SMOTE) to address the issue of imbalanced datasets.

We used sensitivity analysis to remove variables with the least impact on classification performance. The final model identifying irradiated samples was built using 8 miRNAs (file S2): miR-126-5p, miR-320d, miR-10b-5p, miR-375, miR-150-5p, miR-122-5p, miR-215-5p and miR-144-5p. The network had six neurons in the hidden layer and performed very well, with AUC ROC of 0.97 (95%CI: 0.89-1.00) in the training set, sensitivity of 91.7% and specificity of 91.7% in the validation set (Fig. 5F). Network differentiating between high vs low dose (file S3) used levels of miR-126-5p, miR-320d, miR-375, miR-150-5p, miR-122-5p, miR-215-5p and miR-144-5p as input, had four neurons in the hidden layer and showed an AUC ROC of 0.98 (95%CI: 0.90-1.00) in the training set and 83.3% sensitivity with 91.7% specificity in the validation set (Fig. 5F). The diagnostic protocol for the prediction of radiation exposure based on the serum miRNA expression and for the differentiation between high and low dose exposure is presented in Fig. 5G

## DISCUSSION

We demonstrated that a panel of several miRNAs can be used to identify humans exposed to ionizing radiation and differentiate whether the received dose will be high enough to cause life-threatening bone marrow failure. Most of the miRNAs that constitute this signature show evolutionarily conserved patterns of change, confirming a shared regulatory mechanism leading to alterations of their serum levels after exposure.

Using sera collected at several timepoints from patients undergoing TBI as a component of conditioning for HSCT, we were able to reliably evaluate dose-dependent patterns in the expression of radiation-induced miRNAs. We have identified a signature of radiation-induced miRNAs in human serum which allows clear separation of irradiated and non-irradiated patients. We have also pinpointed 7 miRNAs (miR-150-5p, miR-122-5p, miR-122b-3p, miR-574-5p, miR-144-5p, miR-126-5p and miR-200c-3p) differentially expressed in both the low and high dose setting. This moderate overlap is expected, given the genetic heterogeneity of patients and intra-individual differences in the response to radiation. Importantly, all 7 overlapping miRNAs showed consistent direction of change irrespective of the radiation dose level. In six out of the seven miRNAs, the log2FC was greater in the high dose setting, consistent with a real biological effect. Interestingly, when we collated these results with the data obtained from the analysis of the miRNA cargo of serum-derived exosomes, we revealed four shared miRNAs (miR-150-5p, miR-122-5p, miR-126-5p, miR-122b-3p) with a similar pattern of expression changes. Functional enrichment analysis of the exosome-encapsulated miRNAs suggested that the observed exosomal cargo is involved in phenomena crucial for adaptive response. This observation is in line with the reports suggesting that the radiation-induced bystander effect might be mediated by miRNAs contained inside exosomes [32].

Cross-referencing these results with the data from our previous works allowed us to provide the final evidence of conservation of miRNA-based radiation response patterns between species. Importantly, similarities were observed in the level of both individual miRNAs and miRNA families. However, some discrepancies were also noted – for example, miR-133b and miR-133a, important miRNAs used to build classifiers in our earlier study in macaques [19] and with consistent radiation response in mice [18], was absent in human patient sera. These observations have important clinical implications, because miRNAs identified in our study are often evaluated in terms of cancer cell responses to ionizing radiation, [33] and it is possible that these changes are not limited to the malignant cells. Moreover, dysregulated miRNAs have been previously shown to directly target candidate biodosimeter genes such as *ATM* (miR-181 family) [34] or *TP53* (miR-150-5p, miR-375 and miR-30 family) [35].

We also analyzed the cell and tissue origin of miRNAs consistently dysregulated across species and radiation exposure doses, using the data from public repositories [15–17]. We found that the most common putative tissue sources for differentially expressed miRNAs were hematopoietic and endothelial cells, which is in line with observations of Ostheim et al [36]. However, we have also identified other sources of radiation-inducible miRNAs which should be taken into account, e.g. hepatocytes or neuronal stem cells.

Regardless of the exact underlying mechanisms regulating miRNA expression or their tissue sources, the patterns of radiation-induced changes in miRNA serum levels allowed us to translate them into applicable diagnostic tests. A logistic regression classifier enabled efficient classification of the irradiated- and non-irradiated samples in both low and high dose settings. However, to produce a test that can be more rapidly developed for use at the point-of-care, we needed to scale our models to the qPCR technique, which is able to provide results in less than an hour allowing prompt decisions [37]. Based on the premise that a single reference miRNA is statistically worse than a combination in terms of stability and therefore cannot serve as robust normalizer [38], we evaluated all possible pairs, which allowed us to identify the most stable combination of miRNAs – miR-27a-3p and let-7a-5p - whose expression levels were not altered by irradiation. These miRNAs are expressed mostly by fibroblasts and epithelial cells [16]. The release of these miRNAs to serum was not affected by radiation exposure and their expression in all samples markedly exceeded threshold values for detection in qPCR, allowing their use as robust normalizers. After dividing the analysis cohort into training and test set and sensitivity analysis to select variables with greatest impact on classification performance, we designed neural network-based models to identify samples from patients irradiated with any dose and to differentiate between low- and high-dose exposure. This approach yielded classifiers with exceptional performance in a fully independent validation cohort.

The proposed two-tier approach to identifying irradiated individuals and then estimating the radiation dose received is in line with the clinical need of emergency medicine in mass exposure scenarios. Soon after such an event, it is infeasible to determine who received lethal doses of radiation, as typical diagnostic tests are technically difficult and time consuming [39]. A similar concept was recently investigated in a study where the Authors proposed the use of miR-150-5p normalized to one internal reference miRNA to estimate the absorbed radiation dose [40]. The assay, however, was developed and calibrated only for the use in mice, preventing direct assessment of its performance in our dataset of human serum miRNA expression following TBI. As we show using the data from our previous study [18], serum miR-150-5p kinetics in mice and humans differ, in line with differences in radiation sensitivity, body size and the differences in genetic heterogeneity between humans and animal models. Although miR-150-5p enables almost perfect separation of the irradiated from non-irradiated mouse samples, the task is not as straightforward in non-human primates and human patients, posing serious questions regarding the assay translational relevance.

Several classes of biomarkers related to oxidative stress [41], inflammatory and immune response [42], metabolomics [43] and gene expression signatures [44] have been vigorously tested in recent years, with limited success. Multiple studies conducted on animal models [11,18–20,22,40] showed that miRNAs may reliably reflect radiation response and act as functional dosimeters predicting not only the actual dose given but also the biological impact of radiation on the organism [18]. Over the past few years, our group has gradually gathered evidence suggesting that multiple radiation-inducible miRNAs are evolutionarily conserved and that results observed in animals may be translated to humans [18,19]. This notion is also supported by our recent systematic review and meta-analysis which shows that expression levels of miRNAs circulating in body fluids reflect the impact of ionizing radiation regardless of the species, often in a dose-dependent manner [14].

Several limitations of our study should also be noted. Relatively small sample sizes representing heterogenous cohorts of both children and adults, and possible inter-individual differences, might have affected our results. Several samples were rejected after quality control and such potential losses should be also be considered before implementation of the assay. Of note, this issue was observed only in the sequencing part of the study. The sample size for the exosomal cargo analysis was additionally limited, which could potentially restrict our findings, and this part of the study should be perceived as hypotheses-generating and validated in a separate experiment. Despite the use of advanced statistical techniques, we were not able to perfectly discriminate between patients irradiated with low and high doses. Limited sample size might be the possible explanation, but future studies should also focus on the identification of possible additional sources of variability.

Nevertheless, our study shows that a serum miRNA signature reflects radiation exposure and received dose in humans undergoing TBI and may be used as functional biodosimeters for precise identification of people exposed to clinically significant radiation doses.

## MATERIALS AND METHODS

### Study design

This study was designed to identify early serum miRNAs expression changes in human subjects receiving potentially lethal radiation doses and to validate the results of previous similar studies in mice and NHPs. We obtained sera from 25 patients who underwent TBI (2 Gy/fx, total 12 Gy) as a component of a preparative regimen for allogenic HSCT at two radiation oncology centers. The sera were collected at the following timepoints: no irradiation, low dose (2 or 4 Gy; n=25) and high dose (12 Gy, n=9). We profiled miRNA expression in these samples using next-generation sequencing (miRNA-seq). Additionally, we isolated exosomes from paired serum samples of 6 patients collected after exposure to 0 Gy and 12 Gy and performed miRNA-seq on their contents. Following quality control, we performed differential expression testing and constructed logistic regression models of pre-vs post-irradiation miRNA expression profiles. We used publicly available data and bioinformatic tools to identify putative tissue sources and biologic processes associated with top miRNAs with consistent changes in humans, NHPs and mice undergoing TBI.

### Patients

Subjects who were 18 years of age or greater, receiving TBI for any indication, and seen in the Brigham and Women’s Hospital (BWH)’s Department of Radiation Oncology between August 2017 and June 2019 were selected for inclusion in one cohort. The second cohort included pediatric patients receiving TBI for any indication and seen in the Department of Paediatric Bone Marrow Transplantation, Oncology and Hematology, Wroclaw Medical University (WMU), Poland between September 2018 and March 2020.

### Total Body Irradiation procedure

Among the 16 subjects included in this analysis from BWH, 6 subjects received 2 Gy fractions BID over 72 hours to a total dose of 12 Gy, and 10 subjects received a single 2 Gy fraction, administered using a dedicated commercial Cobalt-60 TBI unit. Both radiation protocols are standard treatment for patients at the center. All subjects included in the analysis from WMU received 2 Gy fractions over 72 hours to a total dose of 12 Gy.

### Blood sample collection

Blood samples were collected at pre-specified timepoints via venipuncture. Samples were centrifuged and frozen to -80°C within 4 hours of collection. This work was conducted under Partners IRB approved protocol 2016P001582 (BWH Radiation Oncology All-Department Biorepository to Accelerate New Discoveries, i.e., BROADBAND) and under the approval of Institutional Review Board at Medical University of Lodz (RNN/210/16/KE).

### miRNA profiling by miRNA sequencing

Exosomes were isolated from selected serum samples using the Qiagen miRCURY Exosome Serum/Plasma Kit. Total RNA was isolated using the Qiagen miRNeasy Serum/Plasma Kit. After RNA extraction, small RNA-seq (next generation sequencing) indexed sequencing libraries were prepared from 200 ng total RNA according to QIAseq miRNA Library Kit Handbook (version 11/2016). Sequencing was performed using Illumina NextSeq 550 and 75 bp single-end reads were obtained.

After adapter trimming and quality filtration (on raw fastq files, using fastp and with Phred quality score≥30)[45], unique molecular indexes (UMI) were extracted, and reads were deduplicated with UMI tools[46]. Reads were mapped to miRbase (version 22)[47–52] using bowtie[53] and allowing for 1 mismatch. Resulting bam files were sorted and indexed using samtools[54]. Raw counts were normalized to transcripts per million (TPM), without normalization of library sizes.

### miRNA profiling by qPCR

Selected miRNAs that differed depending on exposure, dose and showed an evolutionarily-conserved pattern of changes were profiled by qPCR with Exiqon (Vedbæk, Denmark) LNA-containing miRNA-specific probes. Briefly, miRNAs were polyadenylated and reverse transcribed into cDNA in a single reaction step, then transferred to pre-loaded plates of primers using a pipetting robot. Amplification was performed on a Roche Lightcycler 480 (Roche, Basel, Switzerland). Amplification quality was determined by generating melting curves; reactions with low efficiency or multiple peaks on the melting curve were discarded. Spike-in positive controls for reverse transcription, qPCR reaction and no template negative controls were included. Minimum detection values for qPCR were established at 37 cycles; miRNAs with no amplification before that number of qPCR cycles were assumed to have their expression undetectable, and a quantification cycle (Cq) value of 37 was imputed as a substitute value. miRNAs with more than 10 observations missing across the dataset were excluded from the analysis. Background filtered and normalized data for samples used to build the model appear in the Data file S1 in accordance with Minimum Information for Publication of Quantitative Real-Time PCR Experiments (MIQE) Guidelines[55].

### Statistical analysis

#### Preprocessing of profiling data

miRNA raw counts were converted to TPM and log10(TPM+0.001)-transformed. The dataset was filtered to samples with at least 350 miRNAs with non-zero reads. Principal Component Analysis (PCA) of all miRNAs detectable in all remaining samples and heatmaps with hierarchical clustering were then used to visually inspect the data, identify and exclude potential outliers. For differential expression testing, paired samples of those identified as having low number of detected miRNAs or classified as outliers were also removed.

### Statistical analysis – miRNA sequencing experiment

For paired analyses, miRNAs were filtered to those with non-zero expression in at least one sample in a pair, in all pairs of samples. Differential expression testing was performed using T.TEST function in Microsoft Excel (paired, two-sided) in the low dose (N=22 pairs), high dose (N=7 pairs) and high dose exosome (N=6 pairs) settings. False discovery rate (FDR) was estimated for each miRNA using Benjamini-Hochberg method. miRNAs with FDR<0.05 in the low dose and FDR<0.25 in the high dose and high dose exosome settings were considered as significantly differently expressed. For the low/high dose comparison and comparisons between human, NHP and mouse studies, all miRNAs with p<0.05 were considered. Log2 fold changes (log2FC) were calculated as the differences in average expression of log2-transformed values before and after irradiation. Correlations of log2FC were performed using Pearson’s correlation test. Hierarchical clustering was performed using Euclidean distance as distance metric and average linkage, with MultiExperiment Viewer (MeV). For the joint low and high dose analysis, we utilized samples from the 9 patients with paired measurements following exposure to 0, 4 and 12 Gy; 2 of them were removed from the analysis due to low number of expressed miRNAs in baseline samples (baseline from pairs 4 and 7; see Supplementary Figure 3). miRNAs were filtered to those with non-zero expression in all three samples from the same patient. To test whether the same set of miRNAs can predict radiation exposure in mice, macaques and humans, we used data from Acharya et al.[18] and Fendler et al.[19] and performed logistic regression limiting the set of miRNAs to those overlapping between species. Similar to the original publication, low dose was defined as 2 Gy in the mouse experiment and high dose was defined as 6.5 or 8 Gy. The regression models were created using 4-fold cross-validation and for each model diagnostic accuracy of the model was reported for the cross-validation by calculating the area under the ROC curve (AUC ROC) with 95% confidence interval. Goodness of fit was evaluated with Hosmer-Lemeshow test. All of the tests were two-sided.

### Functional enrichment analysis

We performed functional enrichment analysis to identify putative biological processes and pathways associated with miRNAs differentially expressed at p<0.05 in exosomes isolated from sera of patients following a total dose of 12 Gy. To this end, we used FunRich - Functional Enrichment analysis tool (http://www.funrich.org)[56] and conducted the analysis in following categories: Molecular Function, Biological Process and Reactome Pathways. For this analysis, we used less stringent filtering variant (miRNAs with non-zero expression in at least one sample in a pair, in >half of pairs; N=565).

### Identifying tissue-specific miRNAs

To identify the source of the radiation-induced miRNAs in serum, we used data regarding the tissue-specific miRNA expression in human from a study by Ludwig et al.[15], tissue-specific expression in human and mouse by Landgraf et al.[17] and data regarding cell type-specific miRNA expression in human and mouse from de Rie et al.[16]. We verified the data regarding high-confidence set of miRNAs and radiation-induced miRNA families consistent across species. miRNAs were considered tissue-specific based on the definition by Ludwig et al., i.e. if their tissue specificity index (TSI) was greater than or equal to 0.85. miRNAs were considered cell-type specific if their Cell type specificity index (CTSI) was greater than 0.9 and the log2 fold ratio of their expression for the first most enriched in cell ontology cluster was greater than 5.

### Normalizer selection

We used miRNA expression of all miRNAs with non-zero counts measured by RNA sequencing as input for NormiRazor tool [38]. The combination with best stability ranking was miR-27a-3p and let-7a-5p. Normalizer stability, i.e. mean miR-27a-3p and let-7a-5p expression compared with mean of all miRNAs evaluated by qPCR, besides normalizers, is presented in Fig. S7A-B.

### Statistical analysis – qPCR experiment

For the validation group, miRNA expression measured by qPCR was normalized to the mean expression of miR-27a-3p and let-7a-5p. Differential expression testing in the low- and high-dose setting was performed using T.TEST function in Microsoft Excel. Correlations of log2FC were performed using Pearson’s correlation test. Hierarchical clustering with Euclidean distance as distance metric and average linkage was used for visualization. Neural networks were trained, 50 thousand at a time, using a grid search algorithm with a combination of linear, logistic, hyperbolic tangens or exponential linking functions. From the analysis set, 14 samples: 7 irradiated ones and 7 non-irradiated ones were randomly selected to a test set for learning optimization. The remaining 19 irradiated samples and 26 nonirradiated ones formed the network training set. To maintain balance of both classes during the learning procedure, we used a Synthetic Minority Oversampling Technique (SMOTE)[57] to balance the training set to a 26:26 case ratio. The secondary set of 12 patients collected from by the Wroclaw center was used as an independent validation set. miRNAs with less than 5 missing values were used to train the neural networks; among the remaining miRNAs, only miR-375 had one missing value and it was filled with the minimum expression value. Sensitivity analysis was used after each training iteration to remove variables with the least impact on classification performance until no further trimming of input variables was possible. The number of neurons in the hidden layer was iteratively optimized from (n=9 variables)/3 to (n=9 variables)*1.5 to avoid overfitting. The best performing network on the training and test set with a less than 10% difference in accuracy between the two sets was then deployed on the validation set for differentiating between the irradiated and non-irradiated patients. Afterwards, a second tier network to differentiate between samples collected at high and low doses. Due to a significant asymmetry of the training and test set (9 high dose samples and 25 low dose samples) we reallocated the samples into a training set of 24 samples and a test set of 10 (5 low/high dose samples each) and used SMOTE to balance the training set. Again, sensitivity analysis was used to reduce the number of input variables to the necessary minimum. The validation set, as it already had a balance of 12 high and 12 low doses, was considered as balanced and unmodified. Diagnostic accuracy of the model was evaluated by calculating the AUC in the analysis set and model performance statistics (sensitivity, specificity and accuracy) in the validation set. Reanalysis of data from Fendler et al.[19] was done after normalizing miRNA expression to the same normalizer that was used in the human dataset, i.e. mean expression of miR-27a-3p and let-7a-5p.

## Data Availability

Raw expression data is deposited in GEO database. qPCR and clinical data are added as supplementary files alongside the manuscript.

https://www.ncbi.nlm.nih.gov/geo/query/acc.cgi?acc=GSE174705

## Data Availability

Data from the miRNA sequencing experiment have been deposited in Gene Expression Omnibus (GEO) and may be found at GEO Accession GSE174705. All other data are available in the main text or the supplementary materials.

## Acknowledgements

This work was supported by Foundation for Polish Science grant First TEAM/2016-2/11 (WF), National Science Centre grant 2019/33/B/NZ5/00536 (WF) and National Science Center grant 2018/29/N/NZ5/02422 (KS). BT gratefully acknowledges financial support provided by the Polish National Agency for Academic Exchange (the Walczak Programme).

## Supplemental Information titles and legends

Fig. S1. Serum samples quality control for the miRNA-seq based experiment.

Fig. S2. Heatmap presenting the expression of all miRNAs with non-zero counts in all serum samples and Principal Component Analysis.

Fig. S3. Scatter plot with correlation coefficient of the log2FC changes in small dose vs log2FC changes in high dose analysis.

Fig. S4. Exosome samples quality control.

Fig. S5. Scatter plot of the log2FC changes in serum high dose vs log2FC changes in exosome high dose analysis.

Fig. S6. Classification models for distinguishing low- and high-dose exposure.

Fig. S7. Normalizer stability in the qPCR experiment.

Fig. S8. miR-150-5p expression kinetics in mouse and human samples.

Fig. S9. Scatterplots of the log2FC of miRNAs selected for validation in RNAseq and qPCR.

Table S1. Clinical data of patients included in the study for the miRNA sequencing experiment.

Table S2. Clinical data of patients included in the study as qPCR validation cohort.

Data file S1. Background filtered and normalized data for samples from the qPCR experiment.

File S2. Neural network model based on the expression of 8 miRNAs predicting ionizing radiation exposure.

File S3. Neural network model based on the expression of 7 miRNAs predicting ionizing radiation dose.

## REFERENCES

1. Burkle FM, Dallas CE. Developing a Nuclear Global Health Workforce Amid the Increasing Threat of a Nuclear Crisis. Vol. 10, Disaster Medicine and Public Health Preparedness. Cambridge University Press; 2016.

2. Coleman CN, Koerner JF. Biodosimetry: Medicine, Science, and Systems to Support the Medical Decision-Maker Following a Large Scale Nuclear or Radiation Incident. Radiat Prot Dosimetry. 2016; 172: 38–46.

3. Coleman CN, Stone HB, Moulder JE, Pellmar TC. Modulation of Radiation Injury. Science (80-). 2004; 304: 693–4.

4. Singh VK, Seed TM. A review of radiation countermeasures focusing on injury-specific medicinals and regulatory approval status: part I. Radiation sub-syndromes, animal models and FDA-approved countermeasures. Int J Radiat Biol. 2017; 93: 851–69.

5. Bolus NE. Basic review of radiation biology and terminology. J Nucl Med Technol. 2017; 45: 259–64.

6. Moulder JE. Post-irradiation approaches to treatment of radiation injuries in the context of radiological terrorism and radiation accidents: A review. Vol. 80, International Journal of Radiation Biology. Int J Radiat Biol; 2004.

7. Rosen EM, Day R, Singh VK. New Approaches to Radiation Protection. Front Oncol. 2015; 4.

8. Obrador E, Salvador R, Villaescusa JI, Soriano JM, Estrela JM, Montoro A. Radioprotection and radiomitigation: From the bench to clinical practice. Vol. 8, Biomedicines. MDPI AG; 2020.

9. Quiat D, Olson EN. MicroRNAs in cardiovascular disease: From pathogenesis to prevention and treatment. Vol. 123, Journal of Clinical Investigation. American Society for Clinical Investigation; 2013.

10. Cortez MA, Bueso-Ramos C, Ferdin J, Lopez-Berestein G, Sood AK, Calin GA. MicroRNAs in body fluids—the mix of hormones and biomarkers. Nat Rev Clin Oncol. 2011; 8: 467–77.

11. Cui W, Ma J, Wang Y, Biswal S. Plasma miRNA as Biomarkers for Assessment of Total-Body Radiation Exposure Dosimetry. Jagetia GC, Ed. PLoS One. 2011; 6: e22988.

12. Templin T, Paul S, Amundson SA, et al. Radiation-induced micro-RNA expression changes in peripheral blood cells of radiotherapy patients. Int J Radiat Oncol Biol Phys. 2011; 80: 549–57.

13. Mitchell PS, Parkin RK, Kroh EM, et al. Circulating microRNAs as stable blood-based markers for cancer detection. Proc Natl Acad Sci U S A. 2008; 105: 10513–8.

14. Małachowska B, Tomasik B, Stawiski K, Kulkarni S, Guha C, Chowdhury D. Circulating microRNAs as Biomarkers of Radiation Exposure : A Systematic Review and Meta-Analysis. 2020; 106: 390–402.

15. Ludwig N, Leidinger P, Becker K, et al. Distribution of miRNA expression across human tissues. 2016; 44: 3865–77.

16. De Rie D, Abugessaisa I, Alam T, et al. An integrated expression atlas of miRNAs and their promoters in human and mouse. Nat Biotechnol. 2017; 35: 872–8.

17. Landgraf P, Rusu M, Sheridan R, et al. A Mammalian microRNA Expression Atlas Based on Small RNA Library Sequencing. Cell. 2007; 129: 1401–14.

18. Acharya SS, Fendler W, Watson J, et al. Serum microRNAs are early indicators of survival after radiation-induced hematopoietic injury. Sci Transl Med. 2015; 7.

19. Fendler W, Malachowska B, Meghani K, et al. Evolutionarily conserved serum microRNAs predict radiation-induced fatality in nonhuman primates. Sci Transl Med. 2017; 9.

20. Menon N, Rogers CJ, Lukaszewicz AI, et al. Detection of Acute Radiation Sickness: A Feasibility Study in Non-Human Primates Circulating miRNAs for Triage in Radiological Events. Amendola R, Ed. PLoS One. 2016; 11: e0167333.

21. Rohrmann S, Linseisen J, Nöthlings U, et al. Meat and fish consumption and risk of pancreatic cancer: Results from the European Prospective Investigation into Cancer and Nutrition. Int J Cancer. 2013; 132: 617–24.

22. Jacob NK, Cooley J V., Yee TN, et al. Identification of Sensitive Serum microRNA Biomarkers for Radiation Biodosimetry. Camphausen K, Ed. PLoS One. 2013; 8: e57603.

23. Jelonek K, Wojakowska A, Marczak L, et al. Ionizing radiation affects protein composition of exosomes secreted in vitro from head and neck squamous cell carcinoma. Acta Biochim Pol. 2015; 62: 265–72.

24. Mutschelknaus L, Peters C, Winkler K, et al. Exosomes derived from squamous head and neck cancer promote cell survival after ionizing radiation. PLoS One. 2016; 11.

25. Arscott WT, Tandle AT, Zhao S, et al. Ionizing Radiation and Glioblastoma Exosomes: Implications in Tumor Biology and Cell Migration. Transl Oncol. 2013; 6: 638–IN6.

26. Le M, Fernandez-Palomo C, McNeill FE, Seymour CB, Rainbow AJ, Mothersill CE. Exosomes are released by bystander cells exposed to radiation-induced biophoton signals: Reconciling the mechanisms mediating the bystander effect. PLoS One. 2017; 12.

27. Hoshino A, Costa-Silva B, Shen T-L, et al. Tumour exosome integrins determine organotropic metastasis. Nature. 2015; 527: 329–35.

28. Lotvall J, Valadi H. Cell to cell signalling via exosomes through esRNA. Cell Adh Migr. 1: 156–8.

29. Miksa M, Wu R, Dong W, et al. Immature Dendritic Cell-Derived Exosomes Rescue Septic Animals Via Milk Fat Globule Epidermal Growth Factor VIII. J Immunol. 2009; 183: 5983–90.

30. Simeoli R, Montague K, Jones HR, et al. Exosomal cargo including microRNA regulates sensory neuron to macrophage communication after nerve trauma. Nat Commun. 2017; 8.

31. Elias KM, Fendler W, Stawiski K, et al. Diagnostic potential for a serum miRNA neural network for detection of ovarian cancer. Elife. 2017; 6.

32. Du Y, Du S, Liu L, et al. Radiation-Induced Bystander Effect can be Transmitted through Exosomes Using miRNAs as Effector Molecules. Vol. 194, Radiation Research. Radiation Research Society; 2020.

33. Czochor JR, Glazer PM. MicroRNAs in cancer cell response to ionizing radiation. Vol. 21, Antioxidants and Redox Signaling. Mary Ann Liebert Inc.; 2014.

34. Rezaeian AH, Khanbabaei H, Calin GA. Therapeutic potential of the miRNA–ATM axis in the management of tumor radioresistance. Vol. 80, Cancer Research. American Association for Cancer Research Inc.; 2020.

35. Feng Z, Zhang C, Wu R, Hu W. Tumor suppressor p53 meets microRNAs. J Mol Cell Biol. 2011; 3: 44–50.

36. Ostheim P, Haupt J, Herodin F, et al. MiRNA Expression Patterns Differ by Total-or Partial-Body Radiation Exposure in Baboons. Radiat Res. 2019; 192: 579–88.

37. Maignan M, Viglino D, Hablot M, et al. Diagnostic accuracy of a rapid RT-PCR assay for point-of-care detection of influenza A/B virus at emergency department admission: A prospective evaluation during the 2017/2018 influenza season. PLoS One. 2019; 14.

38. Grabia S, Smyczynska U, Pagacz K, Fendler W. NormiRazor : tool applying GPU-accelerated computing for determination of internal references in microRNA transcription studies. 2020; 1–16.

39. Chaudhry MA. Biomarkers for human radiation exposure. Vol. 15, Journal of Biomedical Science. J Biomed Sci; 2008.

40. Yadav M, Bhayana S, Liu J, et al. Two-miRNA-based finger-stick assay for estimation of absorbed ionizing radiation dose. Sci Transl Med. 2020; 12.

41. Marrocco I, Altieri F, Peluso I. Measurement and Clinical Significance of Biomarkers of Oxidative Stress in Humans. Vol. 2017, Oxidative Medicine and Cellular Longevity. Hindawi Limited; 2017.

42. Schaue D, Kachikwu EL, McBride WH. Cytokines in Radiobiological Responses: A Review. Radiat Res. 2012; 178: 505–23.

43. Roh C. Metabolomics in radiation-induced biological dosimetry: A mini-review and a polyamine study. Vol. 8, Biomolecules. MDPI AG; 2018.

44. Lu TP, Hsu YY, Lai LC, Tsai MH, Chuang EY. Identification of gene expression biomarkers for predicting radiation exposure. Sci Rep. 2014; 4.

45. Chen S, Zhou Y, Chen Y, Gu J. Fastp: An ultra-fast all-in-one FASTQ preprocessor. In: Bioinformatics. Oxford University Press; 2018.

46. Smith T, Heger A, Sudbery I. UMI-tools: Modeling sequencing errors in Unique Molecular Identifiers to improve quantification accuracy. Genome Res. 2017; 27: 491–9.

47. Kozomara A, Birgaoanu M, Griffiths-Jones S. MiRBase: From microRNA sequences to function. Nucleic Acids Res. 2019; 47: D155–62.

48. Kozomara A, Griffiths-Jones S. MiRBase: Annotating high confidence microRNAs using deep sequencing data. Nucleic Acids Res. 2014; 42: 68–73.

49. Kozomara A, Griffiths-Jones S. MiRBase: Integrating microRNA annotation and deep-sequencing data. Nucleic Acids Res. 2011; 39.

50. Griffiths-Jones S, Saini HK, Van Dongen S, Enright AJ. miRBase: Tools for microRNA genomics. Nucleic Acids Res [Internet]. 2008 [cited 29 January 2021]; 36: D154–8. Available at: https://academic.oup.com/nar/article/36/suppl_1/D154/2507930

51. Griffiths-Jones S, Grocock RJ, van Dongen S, Bateman A, Enright AJ. miRBase: microRNA sequences, targets and gene nomenclature. Nucleic Acids Res [Internet]. 2006 [cited 29 January 2021]; 34: D140–4. Available at: http://microrna.

52. Griffiths-Jones S. The microRNA registry. Nucleic Acids Res [Internet]. 2004 [cited 29 January 2021]; 32: D109–11. Available at: https://academic.oup.com/nar/article/32/suppl_1/D109/2505171

53. Langmead B, Trapnell C, Pop M, Salzberg SL. Ultrafast and memory-efficient alignment of short DNA sequences to the human genome. Genome Biol. 2009; 10.

54. Li H, Handsaker B, Wysoker A, et al. The Sequence Alignment/Map format and SAMtools. Bioinformatics. 2009; 25: 2078–9.

55. Bustin SA, Benes V, Garson JA, et al. The MIQE guidelines: Minimum information for publication of quantitative real-time PCR experiments. Clin Chem. 2009; 55: 611–22.

56. Pathan M, Keerthikumar S, Ang CS, et al. FunRich: An open access standalone functional enrichment and interaction network analysis tool. Proteomics. 2015; 15: 2597–601.

57. Chawla N V., Bowyer KW, Hall LO, Kegelmeyer WP. SMOTE: Synthetic minority over-sampling technique. J Artif Intell Res [Internet]. 2002 [cited 7 March 2021]; 16: 321–57. Available at: https://www.jair.org/index.php/jair/article/view/10302

