## Supplementary Material for "Serum miRNA-based signature indicates radiation exposure and dose in humans: a multicenter diagnostic biomarker study"

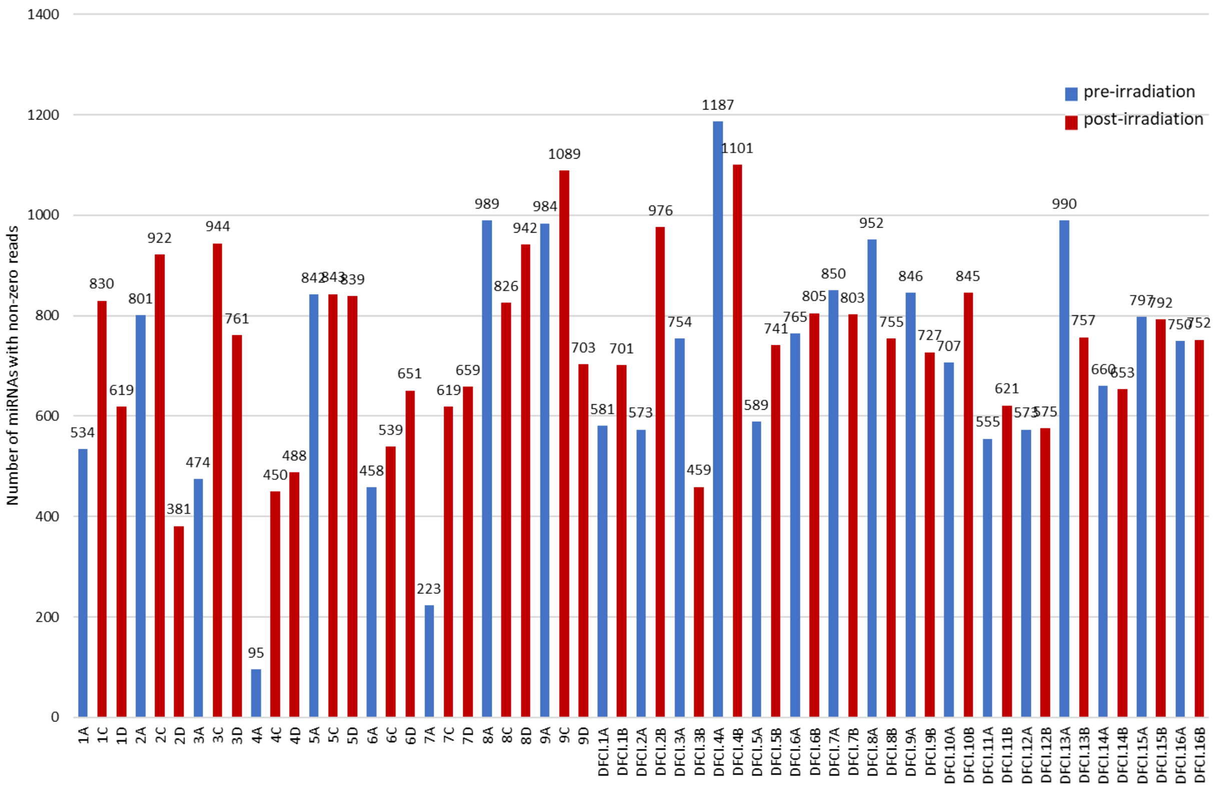


Fig. S1. Serum samples quality control for the microRNA-seq based experiment. Samples with less than 350 miRNAs with non-zero reads detected (4A and 7A) and respective paired samples were removed from the analysis.

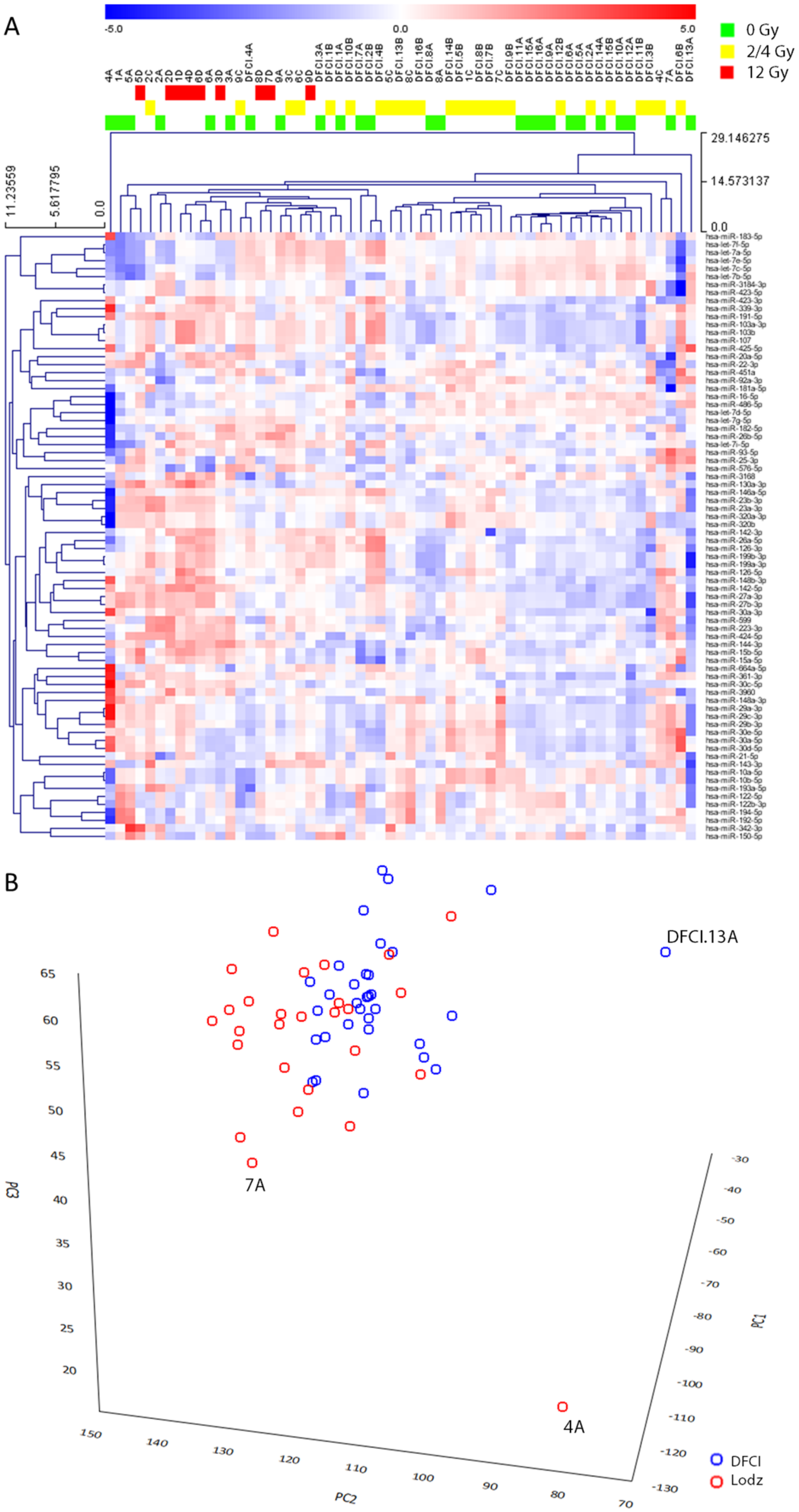
A
-5.0
0.0
5.0
0 Gy
2/4 Gy
12 Gy
29.146275
14.573137
0.0
11.23559
5.617795
0.0
B
DFCI.13A
7A
4A
PC1
PC2
PC3
DFCI
Lodz

Fig. S2. (A) Heatmap presenting the expression of all miRNAs with non-zero counts in all serum samples (B) Principal Component Analysis (PCA) creating using expression of these miRNAs. Besides samples 4A and 7A which had <350 miRNAs with non-zero reads, sample DFCI.13A and respective paired samples were excluded from the analysis as outliers.

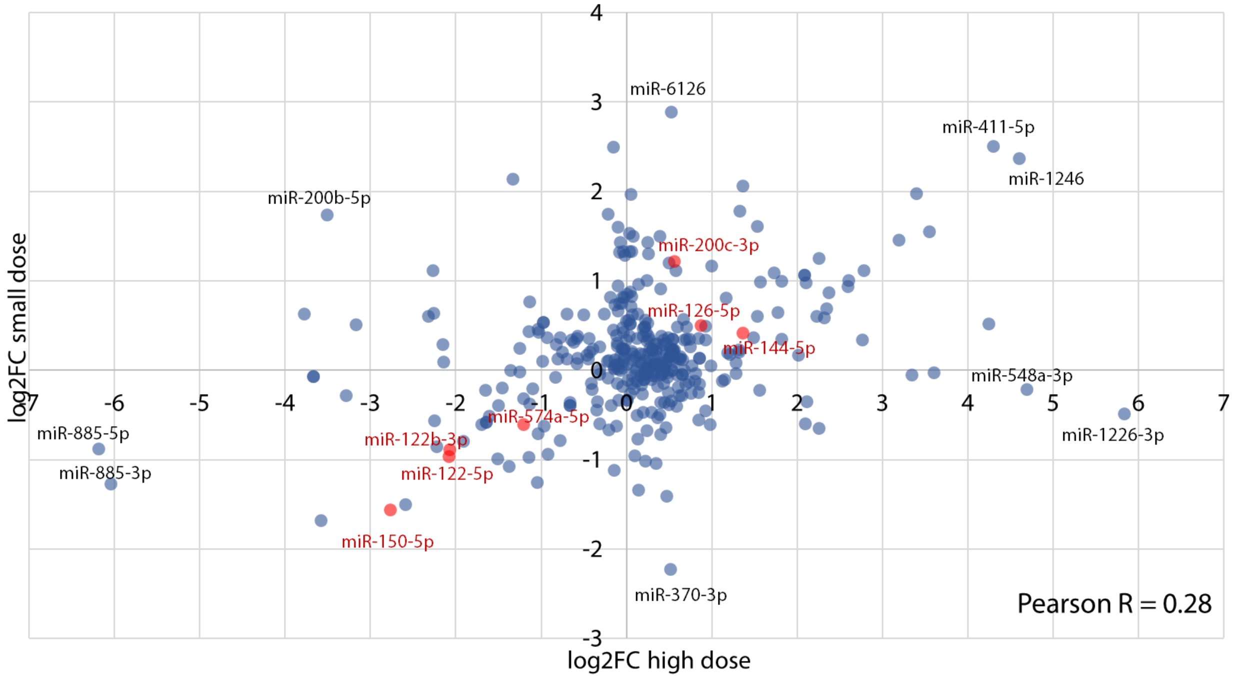


Fig. S3. Scatter plot with correlation coefficient of the log2FC values in the small dose comparison vs log2FC values in the high dose comparison from miRNA sequencing experiment. miRNAs with consistently significant changes in both comparisons ($p<0.05$) are marked with red.

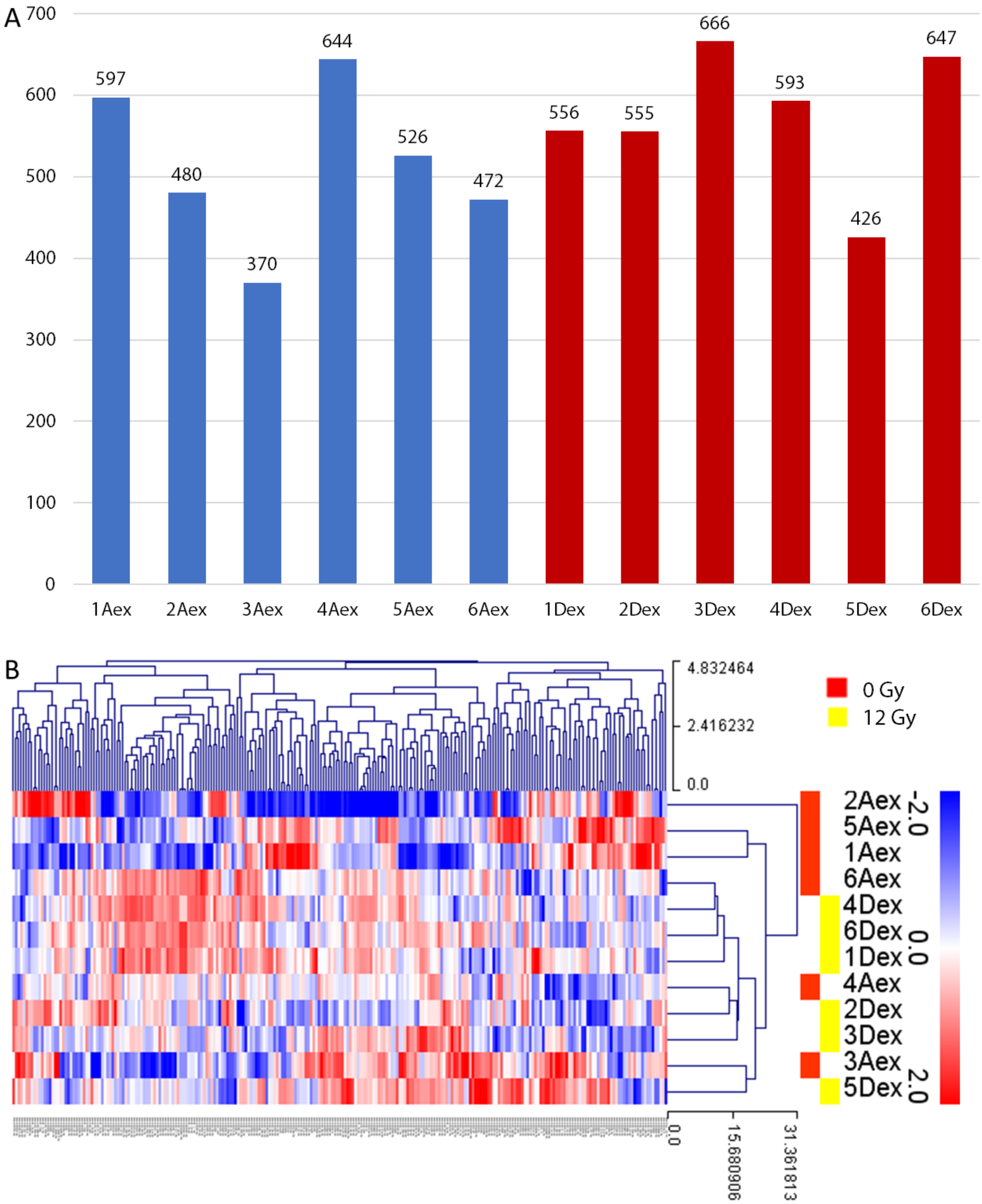


Fig. S4. (A) Exosome samples quality control. None of the samples had <350 miRNAs with non-zero expression. (B) Heatmap presenting the expression of all miRNAs with non-zero counts in all exosome samples.

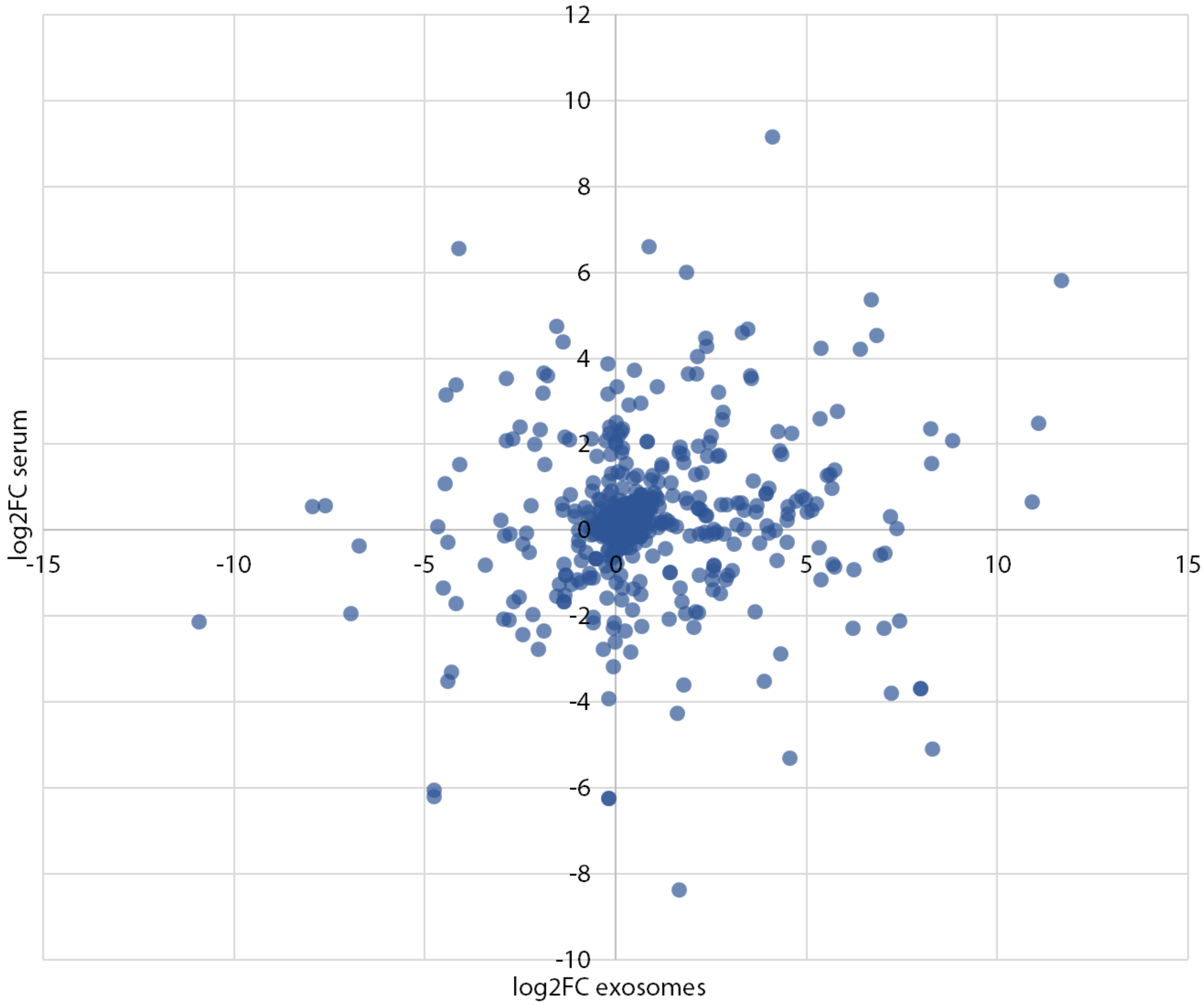


Fig. S5. Scatter plot of the log2FC changes in serum vs log2FC changes in exosomes between samples at baseline and after exposure to 12 Gy.

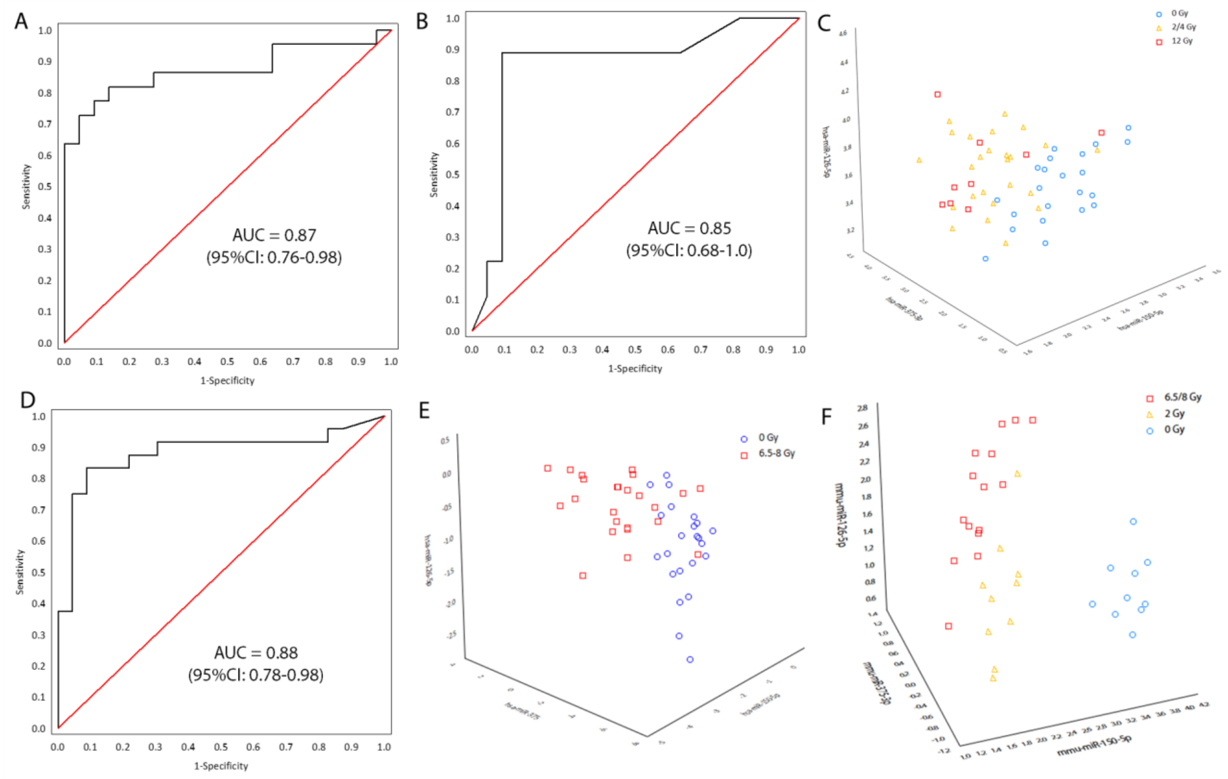


Fig. S6. Classification models for distinguishing low- and high-dose exposure. (A) Performance of a low-dose exposure model in humans. AUROC calculated for a 4-fold cross-validation of a logistic regression model based on expression of miR-150-5p, miR-126-5p and miR-375-3p. (B) Performance of a classification model for detecting high dose exposure in humans using miR—150 and miR-126. AUROC calculated for a 4-fold cross validation procedure. (C) Scatterplot of three miRNAs used in the classifier (miR-150-5p, miR-126-5p and miR-375) in human samples. (D) Performance of a logistic regression model for detecting radiation exposure in macaques using the same 3 miRNAs as in panel A. (E) Scatterplot of three miRNAs used in the classifier (miR-150-5p, miR-126-5p and miR-375) in macaque samples. (F) Scatterplot of three miRNAs used in the classifier (miR-150-5p, miR-126-5p and miR-375) in mouse samples. Performance of both classification models (low- and high-dose exposure) using miRNAs from panels A-C yielded AUROCs of 1.00 with perfect separation of samples.

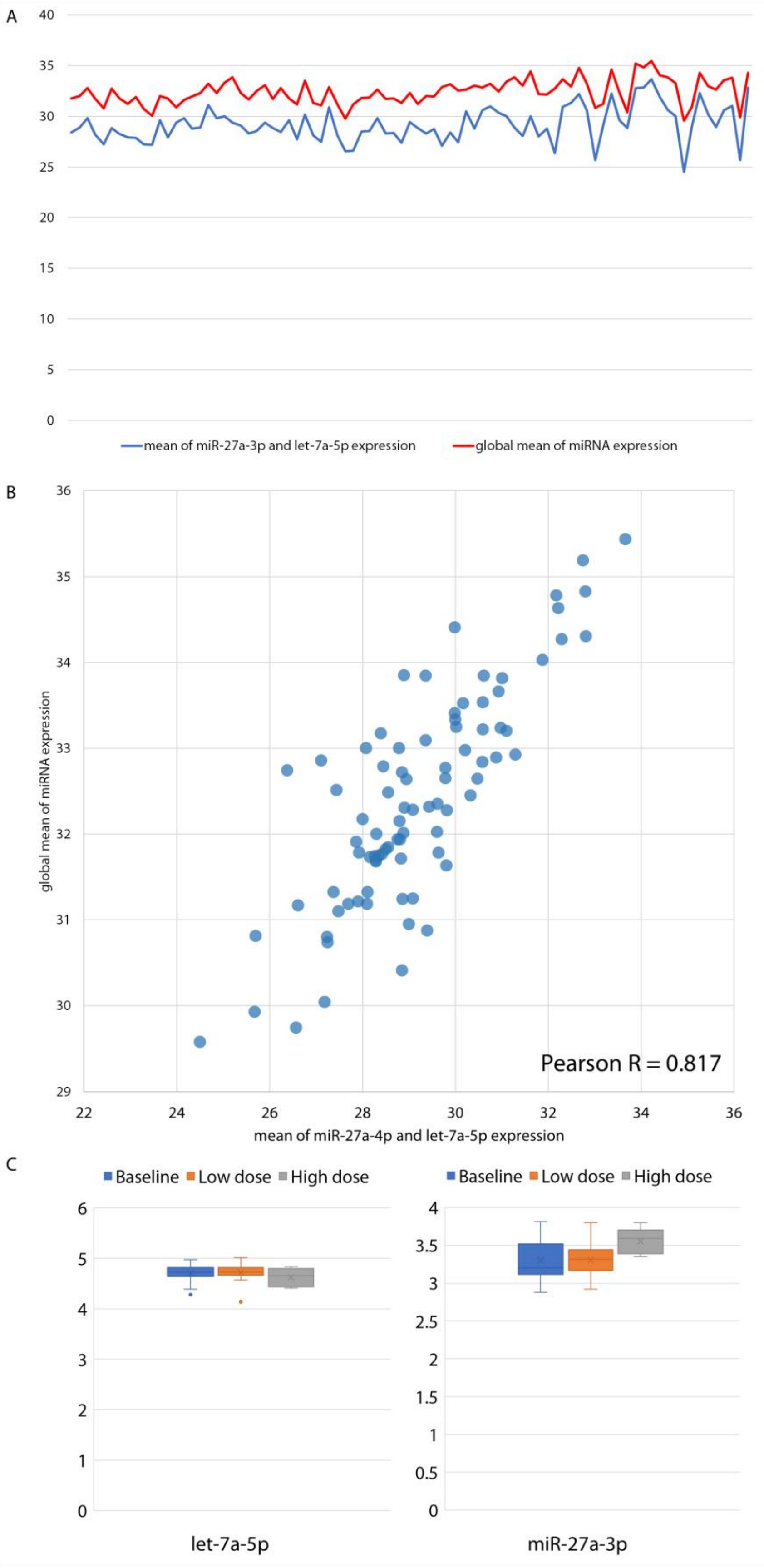
A
40
35
30
25
20
15
10
5
0
mean of miR-27a-3p and let-7a-5p expression
global mean of miRNA expression
B
36
35
34
33
32
31
30
29
global mean of miRNA expression
Pearson R = 0.817
22
24
26
28
30
32
34
36
mean of miR-27a-4p and let-7a-5p expression
C
Baseline
Low dose
High dose
6
5
4
3
2
1
0
let-7a-5p
Baseline
Low dose
High dose
4
3.5
3
2.5
2
1.5
1
0.5
0
miR-27a-3p

Fig. S7. Normalizer stability in the qPCR experiment. (A) Average expression of miR-27a-3p and let-7a-5p (best normalizer according to normiRazor) and the global mean of miRNA expression (without miR-27a-3p and let-7a-5p) across all samples (0/4/12 Gy) in the qPCR experiment. (B) Scatter plot with correlation coefficient of the average normalizer expression and global mean expression of other miRNAs. (C) let-7a-5p and miR-27a-3p expression at baseline, after low dose and high dose exposure measured by microRNA sequencing.

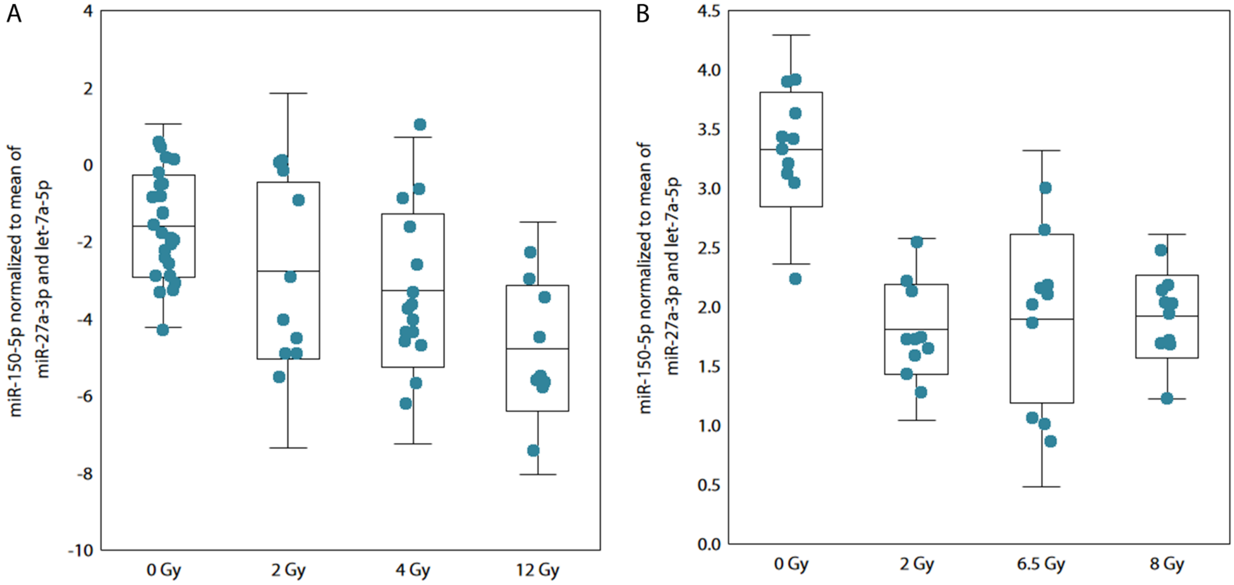


Fig. S8. (A) miR-150-5p expression kinetics in human samples from the qPCR analysis experiment at baseline and after exposure to 2, 4 or 12 Gy. (B) miR-150-5p expression kinetics in mouse samples at baseline and after exposure to 2, 6.5 or 8 Gy, reanalyzed from Acharya et al. (2015)[18]

| Patient number | Center | Sex [M/F] | Primary diagnosis | Age at diagnosis [years range] | Age at transplantation procedure [years range] | status at HSCT | Donor type | Conditioning protocol backbone |
|---|---|---|---|---|---|---|---|---|
| 1 | WMU | M | T-ALL | 6-10 | 6-10 | 1CR | MUD | TBI, VP 60 mg/kg |
| 2 | WMU | M | pre-B ALL | 6-10 | 6-10 | 2CR | MUD | TBI, VP 60 mg/kg |
| 3 | WMU | M | pre-B ALL | 1-5 | 6-10 | 2CR | MSD | TBI, VP 60 mg/kg |
| 4 | WMU | M | T-ALL | 6-10 | 6-10 | 1CR | MSD | TBI, VP 60 mg/kg |
| 5 | WMU | F | NHL hepatopleni | 16-20 | 16-20 | PD | MSD | TBI, VP 60 mg/kg |

| | | | | | | | | |
|---|---|---|---|---|---|---|---|---|
| | | | c T-cell lymphoma | | | | | |
| 6 | WMU | M | T-ALL | 11-15 | 11-15 | 1CR | MSD | TBI, VP 60 mg/kg |
| 7 | WMU | F | T-ALL | 6-10 | 6-10 | 2CR | MUD | TBI, VP 60 mg/kg |
| 8 | WMU | M | ALCL | 6-10 | 11-15 | 2PR | MUD | TBI, FLU 120 mg/m2, MEL 140 mg/m2 |
| 9 | WMU | F | pre-B ALL | 1-5 | 6-10 | 2CR | MUD | TBI, VP 60 mg/kg |
| 10 | BWH | F | ALL (Ph+) | 51-55 | 51-55 | CR | MUD | Myeloablative conditioning (MAC) w/ TBI 1200 cGy in 6 BID fractions, cyclophosphamide (Cy/TBI) |
| 11 | BWH | F | aplastic anemia | 31-35 | 31-35 | UN | MUD | Reduced intensity conditioning (RIC) w/ TBI 200 cGy in 1 fraction, fludarabine, cyclophosphamide, antithymocyte globulin |
| 12 | BWH | M | CLL | 46-50 | 56-60 | PD | MUD | RIC w/ TBI 200 cGy in 1 fraction, fludarabine, cyclophosphamide |
| 13 | BWH | M | acute leukemia with undifferentiated phenotype | 26-30 | 26-30 | CR | MUD | MAC w/ TBI 1200 cGy in 6 BID fractions, cyclophosphamide |

| | | | | | | | | |
|---|---|---|---|---|---|---|---|---|
| 14 | BWH | F | MDS | 61-65 | 61-65 | UN | MFD | RIC w/ TBI 200 cGy in 1 fraction, fludarabine, cyclophosphamide |
| 15 | BWH | M | B-ALL (Ph+) | 61-65 | 61-65 | 3CR | MFD | RIC w/ TBI 200 cGy in 1 fraction, fludarabine, cyclophosphamide |
| 16 | BWH | F | B-ALL | 46-50 | 46-50 | CR | MUD | MAC w/ TBI 1200 cGy in 6 BID fractions, cyclophosphamide |
| 17 | BWH | F | MDS | 66-70 | 71-75 | UN | 1MMUD (mismatched unrelated donor) | RIC w/ TBI 200 cGy in 1 fraction, fludarabine, cyclophosphamide |
| 18 | BWH | M | acute leukemia of ambiguous lineage | 21-25 | 21-25 | CR | MSD | MAC w/ TBI 1200 cGy in 6 BID fractions, cyclophosphamide |
| 19 | BWH | F | B-ALL | 51-55 | 51-55 | CR | MUD | RIC w/ TBI 200 cGy in 1 fraction, fludarabine, cyclophosphamide |
| 20 | BWH | M | Hodgkin lymphoma and chronic myelogenous leukemia | 56-60 | 56-60 | CR | MUD | RIC w/ TBI 200 cGy in 1 fraction, fludarabine, cyclophosphamide |
| 21 | BWH | F | AML | 71-75 | 71-75 | 1CR | double umbilical cord | RIC w/ TBI 200 cGy in 1 fraction, |

| | | | | | | | | |
|---|---|---|---|---|---|---|---|---|
| | | | | | | | blood transplant | fludarabine, cyclophosphamide |
| 22 | BWH | F | B-ALL | 36-40 | 36-40 | 1CR | MSD | MAC w/ TBI 1200 cGy in 6 BID fractions, cyclophosphamide |
| 23 | BWH | F | aplastic anemia | 16-20 | 16-20 | UN | MSD | RIC w/ TBI 200 cGy in 1 fraction, fludarabine, cyclophosphamide, antithymocyte globulin |
| 24 | BWH | M | T-prolymphocytic leukemia | 61-65 | 61-65 | 1CR | 1MMUD | RIC w/ TBI 200 cGy in 1 fraction, fludarabine, cyclophosphamide |
| 25 | BWH | M | Acute biphenotypic leukemia | 41-45 | 41-45 | 1CR | MSD | MAC w/ TBI 1200 cGy in 6 BID fractions, cyclophosphamide |

Table S1. Clinical data of patients included in the study for the miRNA sequencing experiment. ALL – acute lymphoblastic leukemia, AML – acute myelogenous leukemia, CLL – chronic lymphocytic leukemia, CR – complete remission, MDS – myelodysplastic syndrome, MFD – matched family donor, MUD – matched unrelated donor, MMUD – mismatched unrelated donor, MSD – matched sibling donor, NHL – non-Hodgkin lymphoma, PD – progressive disease, TBI – total body irradiation, UN – unknown.

| Patient number | Center | Sex [M/F] | Primary diagnosis | Age at diagnosis [years range] | Age at transplantation procedure [years range] | status at HSCT | Donor type | Conditioning protocol backbone |
|---|---|---|---|---|---|---|---|---|
| 1 | WMU | F | NHL T | 6-10 | 6-10 | 2PR | MUD | TBI, VP 60 mg/kg |

| | | | | | | | | |
|---|---|---|---|---|---|---|---|---|
| 2 | WMU | F | pre-B ALL | 6-10 | 11-15 | 2CR | MUD | TBI, VP 60 mg/kg |
| 3 | WMU | F | T-ALL | 6-10 | 6-10 | 1CR | MSD | TBI, VP 60 mg/kg |
| 4 | WMU | M | T-ALL | 6-10 | 6-10 | 1CR | MSD | TBI, VP 60 mg/kg |
| 5 | WMU | M | T-ALL | 11-15 | 11-15 | 2CR | MUD | TBI, CY 120 mg/kg |
| 6 | WMU | F | pre-B ALL | 6-10 | 6-10 | 1CR | MUD | TBI, VP 60 mg/kg |
| 7 | WMU | M | NHL Burkitt | 6-10 | 6-10 | 2CR | MUD | TBI, VP 60 mg/kg |
| 8 | WMU | F | pre-B ALL | 6-10 | 6-10 | 2CR | MUD | TBI, VP 60 mg/kg |
| 9 | WMU | M | pre-B ALL | 6-10 | 16-20 | 3CR | MSD | TBI, VP 60 mg/kg |
| 10 | WMU | F | pre-B ALL | 6-10 | 11-15 | 1CR | MUD | TBI, VP 60 mg/kg |
| 11 | WMU | M | pre-B ALL | 11-15 | 11-15 | 1CR | MUD | TBI, VP 60 mg/kg |
| 12 | WMU | M | ALCL | 6-10 | 11-15 | 2PR | MUD | TBI, TT 10 mg/kg, VP 40 mg/kg |

Table S2. Clinical data of patients included in the study as qPCR validation cohort. ALL – acute lymphoblastic leukemia, ALCL – Anaplastic large-cell lymphoma, CR – complete remission, MDS – myelodysplastic syndrome, MSD – matched sibling donor, MUD – matched unrelated donor, NHL – non-Hodgkin lymphoma, PR – partial remission, TBI – total body irradiation, UN – unknown.
